# Mpox knowledge and perception in Africa: a systematic review and meta-analysis

**DOI:** 10.64898/2025.12.30.25343224

**Authors:** Fabrice Zobel Lekeumo Cheuyem, Rick Tchamani, Chabeja Achangwa, Ariane Nouko, Jessica Davies, Evariste Mfitumukiza, Constantine Tanywe Asahngwa

## Abstract

**Background:** Mpox, a zoonotic disease long endemic in Africa, has gained renewed global attention due to recent outbreaks. Effective control of the virus relies on public adherence to preventive measures, which is largely influenced by the population’s knowledge and perception. This systematic review and meta-analysis aimed to determine the pooled prevalence of good knowledge and positive perception toward mpox in Africa and to identify associated factors.

**Methods:** Following PRISMA guidelines, a comprehensive search was conducted across multiple relevant databases and grey literature sources. Studies conducted in African countries that assessed knowledge and/or perception of mpox were included. Pooled prevalences with 95% confidence intervals (CIs) were calculated using a random-effects model. Subgroup analyses and meta-regression were performed to explore heterogeneity.

**Results:** The analysis incorporated 38 studies with 23,648 participants from 15 African countries. The pooled prevalence of good mpox knowledge was 43.12% (95% CI: 34.38–52.32), indicating significant gaps in awareness. Knowledge levels varied substantially across subgroups. By participant type, teachers exhibited the highest knowledge (89.43%), followed by medical students (56.02%) and healthcare workers (51.39%), while the general population demonstrated markedly lower knowledge (14.88%). Geographically, Southern Africa had the highest knowledge prevalence (77.92%), whereas Central Africa had the lowest (19.20%). At the country level, South Africa (77.92%) and Kenya (68.31%) recorded the highest levels, while Libya (5.34%) and Somalia (9.68%) had the lowest. The pooled prevalence of a positive perception or attitude toward mpox was 54.22% (95% CI: 44.94–63.21). Pregnant women (81.43%) and healthcare workers (60.32%) reported the most positive perceptions, whereas community health workers showed the least (10.19%). Positive perception was highest in Northern Africa (63.02%) and lowest in Central Africa (7.27%). Individual levels of mpox knowledge and perception were significantly associated with several sociodemographic and non-sociodemographic factors.

**Conclusions:** This study reveals that overall knowledge of mpox across Africa is insufficient, and positive perception is only moderate, with considerable disparities across different populations and regions. The findings underscore an urgent need for targeted educational campaigns, enhanced training for healthcare workers, and context-specific communication strategies to boost awareness, improve attitudes, and strengthen continent-wide outbreak control and preparedness.

## 1. Background

Mpox is a zoonotic disease caused by the monkeypox virus [1]. This virus spreads through direct contact with infected animals or individuals, including through sexual relations or skin-to-skin contact, as well as via respiratory droplets and contaminated objects such as clothing, towels, and bedding [2,3].The infection has garnered increasing international attention due to changes in its epidemiology [4]. Prior to 2022, it was primarily reported in isolated communities in the tropical forests of West and Central Africa [5]. Although this disease has historically been confined to these regions, recent outbreaks have demonstrated its ability to spread beyond endemic areas, posing a significant risk to global public health [6]. Since the beginning of 2025, Africa has recorded 578 deaths attributed to Mpox [7].

The severity of the disease is variable, ranging from mild forms to severe cases, the latter being mainly observed in immunocompromised individuals with potential systemic complications [8–10]. Additionally, the lethality of Mpox differs according to viral clades. On the one hand, clade I is associated with higher mortality, reaching up to 10%, whereas clade II generally has a more favorable prognosis. Nevertheless, recent studies conducted in the Democratic Republic of Congo confirm the persistence of virulent strains and sustained transmission, including new variants (clade Ib), with a significant impact in terms of mortality [11,12].

To address this pandemic, vaccination is an essential pillar of prevention. However, its success is hampered by significant vaccine hesitancy, which is itself influenced by an insufficient level of knowledge about the disease and misperceptions regarding its severity [13,14]. Yet, the foundation for adherence to preventive measures relies on an adequate knowledge and perception of the disease [15–17]. Numerous surveys highlight an insufficient level of knowledge regarding Mpox among various groups, including healthcare professionals and the general public [18–21]. These studies consistently point to gaps concerning modes of transmission, clinical signs, prevention measures, and reliable sources of information [20,22–27].

Several factors have been identified as being associated with poor knowledge of mpox. Insufficient media coverage or limited access to accurate information has been correlated with a lack of awareness about the disease [19,22,28]. Furthermore, occupation and education level are other determining factors; indeed, non-medical personnel, and individuals with lower education levels generally demonstrate lower levels of knowledge [23,29,30]. Likewise, a lack of specific training and clinical exposure is a significant determinant, as noted in studies involving healthcare workers and medical students [27,31]. Moreover, geographical context and the resource levels of healthcare facilities also play a role, with knowledge often being lower in rural areas or less equipped regions [21–23]. Beyond factual knowledge, the perception of risk of the disease itself plays a fundamental role in the adoption of protective behaviors. Despite the severity of this disease, public awareness remains low [20,32], and the perceived risk of infection is limited [25,33].

To date, several studies have been conducted yielding varying level of knowledge and attitude toward the disease which may contribute to the virus’s continued spread across the continent [19,29,34]. Establishing a consolidated evidence base is crucial to address this gap in literature. This systematic review and meta-analysis aim to determine the pooled prevalence of good knowledge and positive perception or attitude toward mpox in Africa and to systematically identify associated factors. The findings are essential for developing targeted interventions, such as tailored communication and vaccination strategies for key populations like healthcare workers and local communities, to control the virus and mitigate its impacts in the continent effectively.

## 2. Methods

### 2.1. Study design, protocol and registration

The PRISMA checklist recommendations were followed in conducting this systematic review and meta-analysis. To ensure process rigor and transparency, the study protocol was properly filed in the International Prospective Register of Systematic Reviews <u>CRD420251133843</u> [35,36].

### 2.2. Eligibility criteria

#### 2.2.1. Inclusion criteria

We included observational and interventional studies that assessed the prevalence of knowledge, perception, or both, of mpox, provided they were fully available, specified the sample size, and presented directly usable data or data that allowed for the calculation of the sought outcomes. There were no timeframe restrictions for this review to capture both historical data on smallpox knowledge and perception and more recent information from mpox outbreaks. Finally, we only considered studies published in either English or French and conducted in African countries.

#### 2.2.2. Exclusion criteria

Studies were excluded for the following reasons: duplication of data, focus outside the scope of our research objectives, non-observational studies (comment, letter to editor, review, other systematic review) and studies conducted outside Africa. Additionally, articles lacking full-text availability were omitted due to insufficient data for analysis or absence of required outcome measures.

### 2.3. Search strategy and information sources

#### 2.3.1. Databases

A comprehensive search strategy was employed to identify all relevant literature. Multiple electronic databases were searched, including PubMed, Scopus, ScienceDirect, Web of Science, CINAHL, and EMBASE. To incorporate research from African scholars, African Journals Online (AJOL) was also consulted. In addition to these databases, we searched for grey literature, which includes unpublished research and preprints. Moreover, manual search was conducted on google scholar and on reference lists of included studies.

#### 2.3.2. Search terms

To capture the relevant literature effectively, we used a comprehensive search strategy that combines Medical Subject Headings (MeSH) terms and Boolean connectors. The search used the following terms and logic: (“Mpox” OR “monkeypox”) AND (“knowledge” OR “perception” OR “awareness” OR “attitude”) AND (“Africa” OR [individual country names, e.g., “Cameroon”, “Democratic Republic of the Congo”]). The final search was completed in February 2025 and updated on August 07, 2025.

### 2.4. Data extraction

Data extraction followed a systematic multi-step procedure. After importing all selected articles into Zotero software version 7.0.16 (Corporation for Digital Scholarship, Fairfax, Virginia, USA), duplicate entries were removed. One investigator (FZLC) extracted key study characteristics using a standardized Microsoft Excel 2016 form, capturing information including the first author’s name, year of study completion, country, study design, participant type, setting, sampling technique, counts of participants with good mpox knowledge and positive attitudes, total sample size, and factors associated with knowledge and perception of mpox. Two additional investigators (RT and CA) independently verified the extracted data for accuracy.

### 2.5. Quality assessment of included studies

Following data extraction, the quality of each included study was independently assessed by two reviewers (FZLC and AN) using the Joanna Briggs Institute (JBI) critical appraisal tool [37]. For cross-sectional studies, the appraisal criteria included: a clear definition of the study population and setting; the use of valid and reliable methods for measuring exposures and outcomes; the identification and management of confounding factors; and the appropriateness of the statistical analyses. Each item was scored as “yes” (1) or “no/unclear” (0). The overall risk of bias was then categorized as low (> 50%), moderate (25–50%), or high (< 25%). Any disagreements between the reviewers were resolved through discussion or by consultation with a third reviewer (CTA).

### 2.6. Study outcome

This analysis focused on two primary outcomes: knowledge and perception or attitude toward mpox. The assessment of participant knowledge was based on definitions provided in the included studies, which characterized “good knowledge” as a demonstrated understanding of key aspects of mpox, including its transmission modes, clinical symptoms, diagnosis, treatment, and prevention [18,19,38]. the assessment of perception or attitude was derived from the original studies’ metrics. A “positive perception or attitude” was defined as confidence in the overall ability to control the epidemic, in the effectiveness of preventive and control measures, and in the perception that health actions are adequate to prevent its spread such as vaccination [18,29,38].

### 2.7. Statistical analysis

The proportions of participants with good knowledge and positive perception were calculated for each study by dividing the number of positive responses by the total sample size and multiplying by 100. To investigate potential sources of heterogeneity, we performed subgroup analyses based on study period, country, setting, participant type, and sampling technique. Countries were categorized according to the WHO African Region classification [39]: Western (Algeria, Ghana, Nigeria), Eastern (Ethiopia, Kenya, Uganda), Southern (South Africa), and Central (Cameroon). Egypt and Morocco were analyzed as a separate Northern African (non-WHO Afro) subgroup. The association between these study-level characteristics and the pooled estimates was further examined using univariable and multivariable meta-regression.

Study heterogeneity was assessed using the Cochrane Q statistic and quantified with the *I²* statistic, with values of 25%, 50%, and 75% representing low, moderate, and high heterogeneity, respectively. A random-effects model was applied for all pooled analyses. We utilized the Generalized Linear Mixed Models (GLMM), coupled with the Probit-Logit Transformation (PLOGIT), which is robust for synthesizing proportional data, including extreme proportions of 0% or 100%, without needing continuity corrections [40]. Statistical significance was defined as a two-sided *p*-value < 0.05. All analyses were performed using R software version 4.4.2 with the ‘meta’ package [41], and maps were created with the MapChart online application [42].

### 2.8. Publication bias and sensitivity analysis

The funnel plot was used to assess potential publication bias. In addition, statistical evaluation was conducted using Egger’s linear regression test and Begg’s rank correlation test, with a p-value < 0.05 indicating potential publication bias. A *p*-valuelll>lll0.05 indicating no statistically significant evidence of publication bias. Sensitivity analysis was conducted by iteratively excluding one study at a time to assess the robustness of the findings.

## 3. Results

The systematic search identified 3,058 records (3,053 from databases and 5 from other sources). After removing 736 duplicates, we screened records by title and abstract and then by full-text. Of these, 38 primary research articles met the eligibility criteria and were included in the meta-analysis [3,13,14,19,21–31,33,34,43–62]. The complete study selection process is detailed in the PRISMA flow diagram (Fig. 1).

**Fig. 1.**
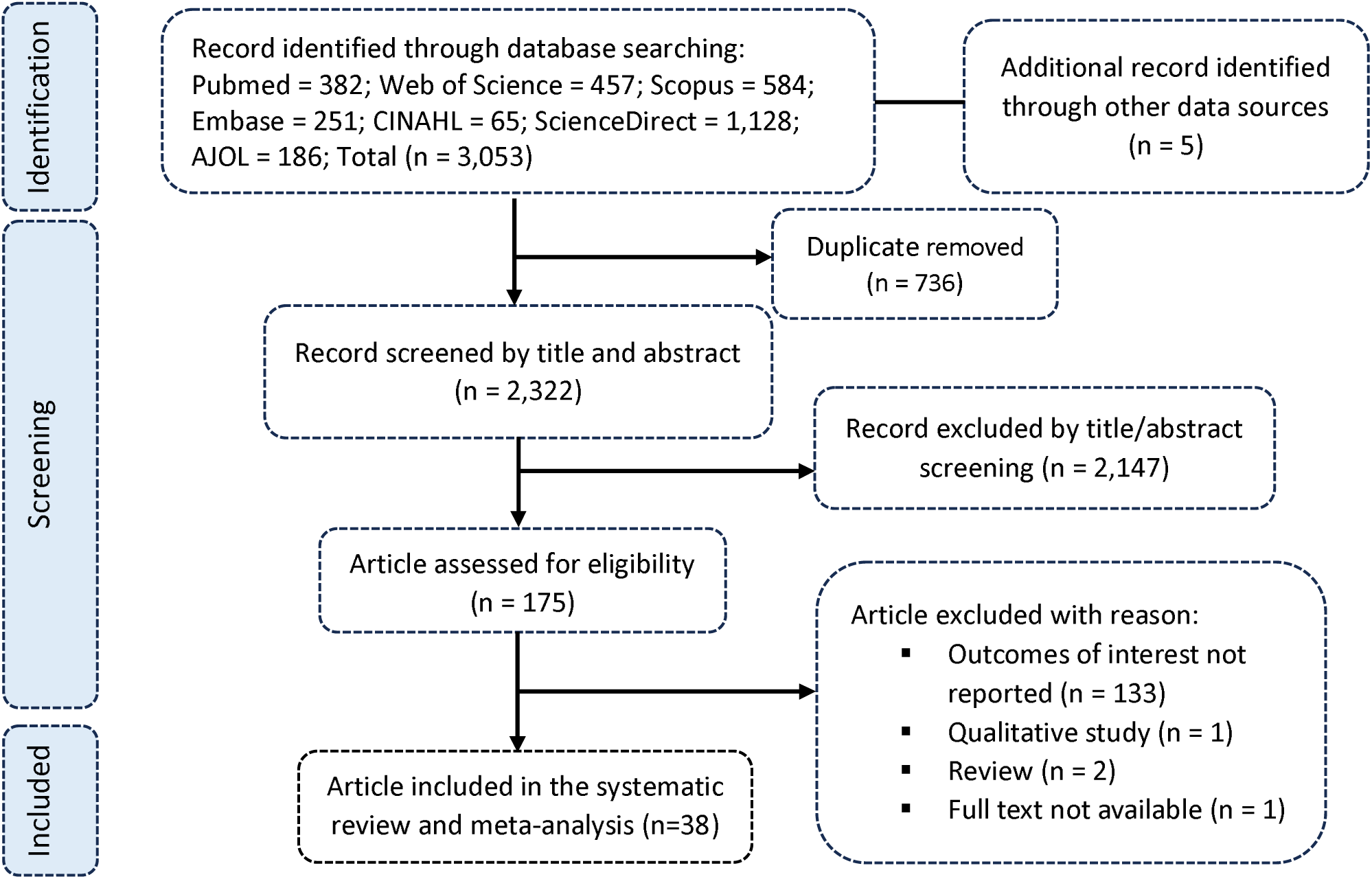
PRISMA diagram flow from study identification to inclusion in the meta-analysis

### 3.1. Studies selection

### 3.2. Characteristics of included studies

This systematic review and meta-analysis incorporated 38 studies comprising a total sample size of 23,648 participants from 15 African countries. The studies were conducted between 2021 and 2025, with the majority (30 studies = 78.9%) conducted in 2022-2023. The included studies represented diverse African regions: West Africa (Nigeria and Senegal); North Africa (Egypt, Algeria, Libya, Morocco, Tunisia); East Africa (Ethiopia, Kenya, Uganda, Rwanda, Somalia, Sudan and Tanzania); Central Africa (Cameroon), Southern Africa (South Africa) and one multicountry study. Studies included were predominantly cross-sectional (36 studies = 94.7%) with two quasi-experimental studies. The settings varied across online platforms (17 studies = 44.7%), healthcare facilities (14 studies = 36.8%), community settings (3 studies = 7.9%), schools (2 studies = 5.2%), and mixed settings (2 studies = 5.2%). Participants were primarily healthcare workers (22 studies = 57.9%), and general population (8 studies = 21.0%), with representation from students, teachers, hotel workers, pregnant women, and community health workers. The sample sizes ranged from 4 to 1,452 participants. The prevalence of good knowledge, reported in 35 studies, ranged from 0% (Morocco general population) to 97.22% (Egyptian teachers). A positive perception or attitude was reported in 20 studies (52.6%), with prevalence ranging from 7.27% (Cameroon general population) to 89.32% (Egyptian healthcare workers). Most studies used non-probabilistic sampling (24 studies = 63.2%). The risk of bias was mostly low (31 studies = 81.6%) (Table 1).

**Table 1.**
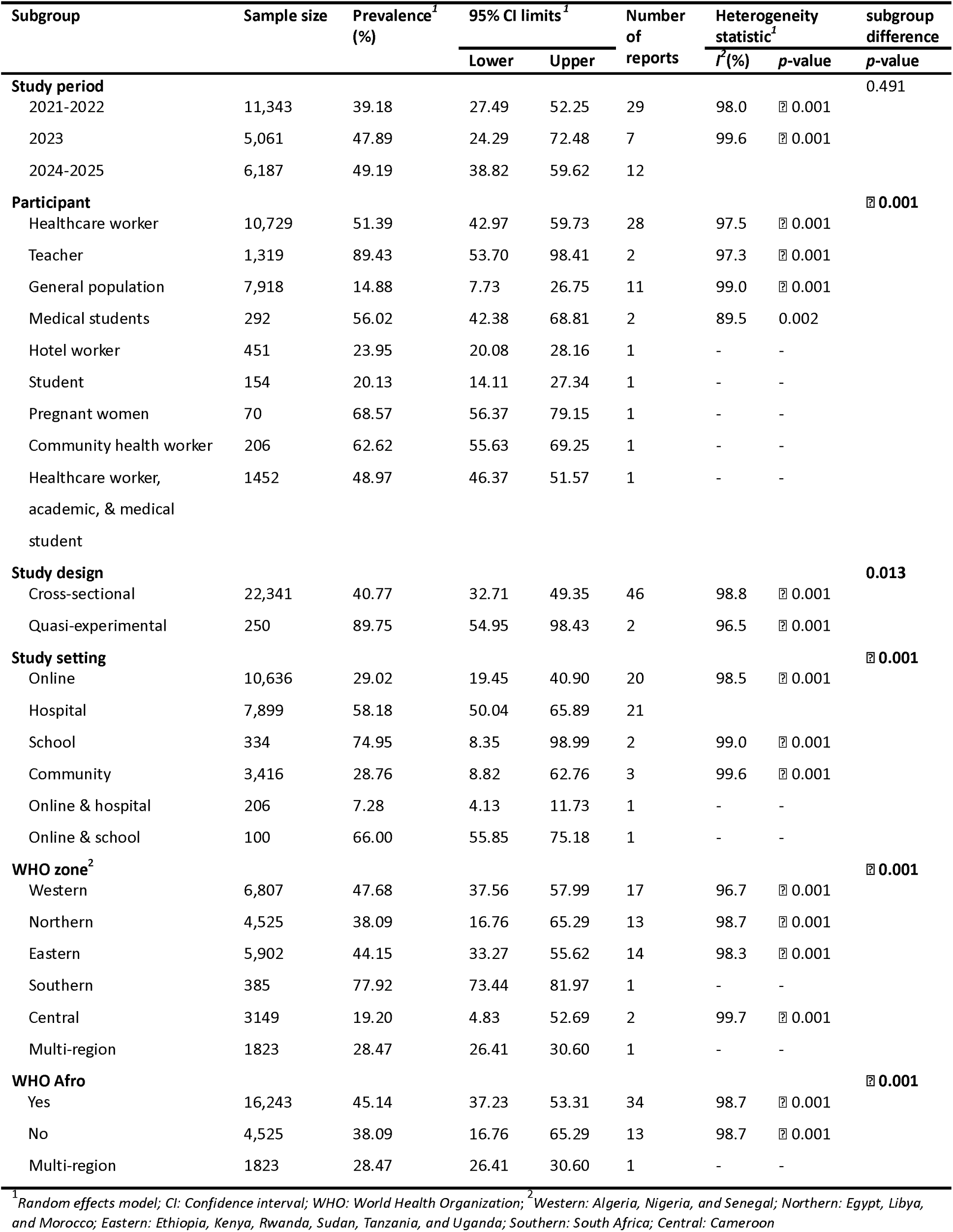
Subgroup meta-analysis of the pooled prevalence estimates of good mpox knowledge in Africa.

### 3.3. Level of knowledge of mpox

The random-effects meta-analysis of 48 country specific reports (n = 22,591) from 35 primary research studies found that the pooled prevalence of good mpox knowledge in Africa was 43.12% (95% CI: 34.38–52.32), with high heterogeneity among studies (*I²* = 98.7%, *p* < 0.001) (Fig. 2).

**Fig. 2.**
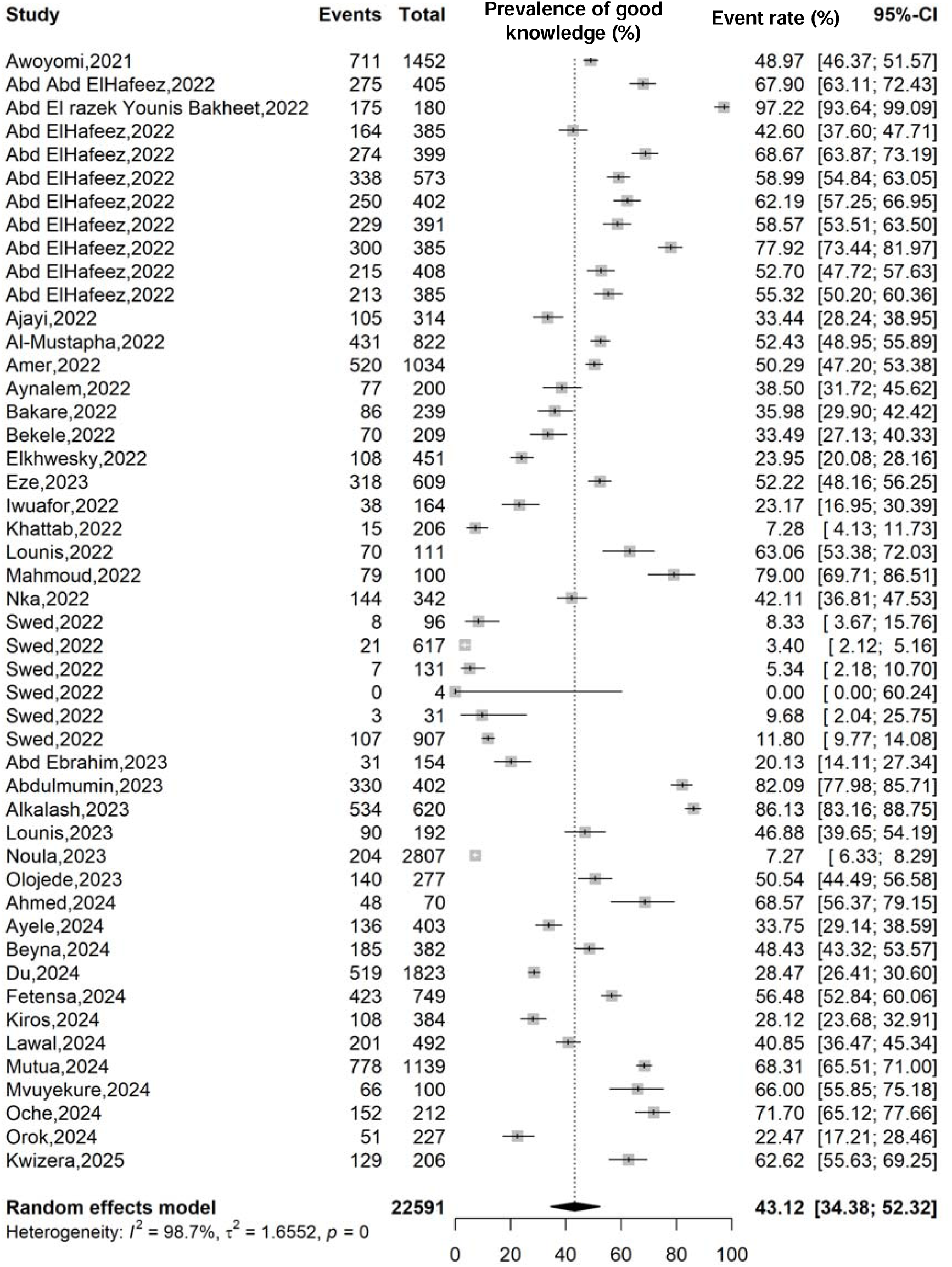
Forest plot displaying the pooled prevalence of good knowledge on mpox in Africa

Subgroup analyses revealed significant disparities in mpox knowledge. Although not statistically significant (*p* = 0.491), the prevalence trend appeared to increase over time from 39.18% (95% CI: 27.49–52.25; n = 11,343; 29 reports) in 2021-2022 [13,19,21,22,25,29–31,43–46,50–53] to 49.19% (95% CI: 38.82–59.62; n = 6,187; 12 reports) in 2024-2025 [3,14,23,24,26,28,33,34,58–60,62]. Conversely, knowledge levels differed significantly by participant type (*p* < 0.001), with teachers exhibiting the highest pooled prevalence (89.43%; 95% CI: 53.70–98.41; n = 1,319; 2 reports) [33,46], followed by Medical students (56.02%; 95% CI: 42.38–68.81; n = 292; 2 reports) [27,28] compared to healthcare workers (51.39%; 95%CI 42.97-69.73; n = 10,729; 28 reports) [3,13,14,21,24–26,29,31,44,45,47,50–55,59,60], and the general population who demonstrated substantially lower knowledge (14.88%; 95% CI: 7.73–26.75; n = 7,918; 11 reports) [19,20,22,23,34,57].

Geographically, knowledge varied significantly across the WHO African regions (*p* < 0.001), with Southern Africa showing the highest prevalence (77.92%; 95% CI: 73.44–81.97; n = 385; 1 report) [29], followed by Western Africa (47.68%; 95% CI: 37.56–57.99; n = 6,807; 17 reports)[19,22,24,26,27,29,43–45,47,50,52,54,57,59] while Central Africa had the lowest (19.20%; 95% CI: 4.83–52.69; n = 3,149; 2 reports) [20,21]. The good mpox knowledge prevalence was significantly (*p* lll 0.001) higher in the WHO Afro region (45.14%; 95%CI: 37.23–53.31; n = 16,243; 34 reports) [3,13,14,19–21,23,24,26–29,31,33,43–45,47,50,52,54,57,59,60,62] compared to non-WHO Afro region (38.09%; 95%CI: 16.76–65.29; n = 4,525; 13 reports)[19,25,29,30,46,51,53,55,56,58] (Table 2 and Additional File 2, Supplementary Fig. 1-8).

**Table 2.** Subgroup meta-analysis of the pooled prevalence estimates of positive mpox perception in Africa.

| Subgroup | Sample size | Prevalence<sup>1</sup><br>(%) | 95% CI limits<sup>1</sup> |  | Number<br>of<br>reports | Heterogeneity<br>statistic<sup>1</sup> |  | Subgroup<br>difference<br>*p*-value |
| --- | --- | --- | --- | --- | --- | --- | --- | --- |
|  |  |  | Lower | Upper |  | *I*<sup>2</sup>(%) | *p*-value |  |
| **Study period** |  |  |  |  |  |  |  | 0.362 |
| 2021-2022 | 7,693 | 57.80 | 49.18 | 65.97 | 17 | 97.9 | ⊠ 0.001 |  |
| 2023 | 3,363 | 29.81 | 8.09 | 67.20 | 3 | 99.7 | ⊠ 0.001 |  |
| 2024-2025 | 2,434 | 55.80 | 37.02 | 73.05 | 8 | 97.4 | ⊠ 0.001 |  |
| **Participant** |  |  |  |  |  |  |  | ⊠ 0.001 |
| Healthcare worker | 8,218 | 60.32 | 53.50 | 66.77 | 21 | 96.1 | ⊠ 0.001 |  |
| General population | 3,210 | 24.45 | 4.31 | 69.90 | 2 | 99.8 | ⊠ 0.001 |  |
| Teacher | 180 | 68.89 | 61.58 | 75.57 | 1 | - | - |  |
| Student | 154 | 29.87 | 22.77 | 37.76 | 1 | - | - |  |
| Pregnant women | 70 | 81.43 | 70.34 | 89.72 | 1 | - | - |  |
| Community health worker | 206 | 10.19 | 6.42 | 15.16 | 1 | - | - |  |
| Healthcare worker,<br>academic, & medical<br>student | 1,432 | 23.69 | 21.53 | 25.96 | 1 | - | - |  |
| **Study design** |  |  |  |  |  |  |  | 0.002 |
| Cross-sectional | 13,240 | 52.43 | 42.87 | 61.82 | 26 | 98.9 | ⊠ 0.001 |  |
| Quasi-experimental | 250 | 74.00 | 63.89 | 82.07 | 2 | 74.2 | 0.049 |  |
| **Study setting** |  |  |  |  |  |  |  | ⊠ 0.001 |
| Online | 2,959 | 52.08 | 34.25 | 69.40 | 5 | 99.0 | ⊠ 0.001 |  |
| Hospital | 6,575 | 59.37 | 52.64 | 65.77 | 17 | 95.7 | ⊠ 0.001 |  |
| School | 334 | 49.33 | 23.57 | 75.45 | 2 | 97.9 | ⊠ 0.001 |  |
| Community | 3,416 | 18.57 | 5.12 | 49.07 | 3 | 99.6 | ⊠ 0.001 |  |
| Online & hospital | 206 | 89.32 | 84.28 | 93.19 | 1 | - | - |  |
| **WHO zone<sup>2</sup>** |  |  |  |  |  |  |  | ⊠ 0.001 |
| Western | 4,160 | 55.50 | 41.23 | 68.91 | 8 | 98.9 | ⊠ 0.001 |  |
| Northern | 3,101 | 63.02 | 48.84 | 75.25 | 8 | 96.1 | ⊠ 0.001 |  |
| Eastern | 3,037 | 50.84 | 37.11 | 64.45 | 9 | 96.3 | ⊠ 0.001 |  |
| Southern | 385 | 61.30 | 56.23 | 66.19 | 1 | - | - |  |
| Central | 2,807 | 7.27 | 6.33 | 8.29 | 1 | - | - |  |
| **WHO Afro** |  |  |  |  |  |  |  | 0.154 |
| Yes | 10,389 | 49.94 | 38.75 | 61.13 | 19 | 99.1 | ⊠ 0.001 |  |
| No | 3,101 | 63.02 | 48.84 | 75.25 | 9 | 96.1 | ⊠ 0.001 |  |

<sup>1</sup> Random effects model; CI: Confidence interval; WHO: World Health Organization; <sup>2</sup> Western: Algeria, Nigeria, and Senegal; Northern: Egypt and Morocco; Eastern: Ethiopia, Sudan, Tanzania, and Uganda; Southern: South Africa; Central: Cameroon

At country level, the lowest prevalence of good mpox knowledge was significantly (*p* < 0.001) observed in Libya (5.34%; 95%CI: 2.18–10.70; n = 131; 1 report) [19] and Somalia (9.68%; 95%CI: 2.04–25.75; n = 31; 1 report) [19]. Conversely, highest prevalences were recorded in South Africa (77.92%; 95%CI: 73.44–81.97; n = 385; 1 report), and Kenya (68.31%; 95%CI: 65.51–71.00; n = 1,139; 1 report) [33], Rwanda (66%; 95%CI: 55.85–75.18; n = 100; 1 report) [28], and Uganda (62.62%; 95%CI: 55.63–69.25; n = 206; 1 report) [62] (Fig. 3).

**Fig. 3.**
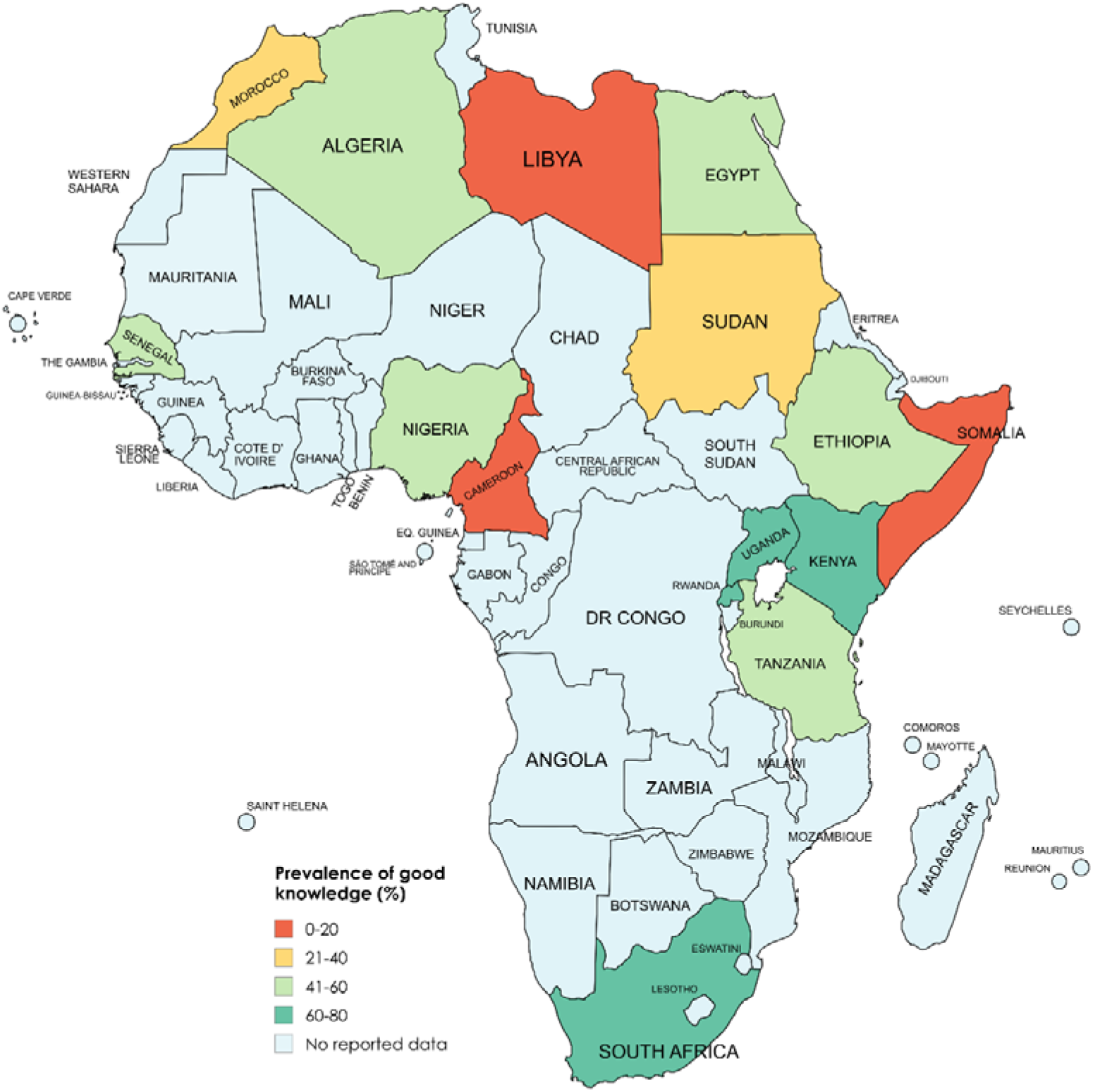
Map of the prevalence of good mpox knowledge across African countries (Additional File 2, Supplementary Fig. 2)

### 3.4. Positive perception toward mpox

The pooled prevalence of positive perception toward mpox among 13,490 total participants from 28 reports was 54.22% (95%CI: 44.94-63.21). A high heterogeneity was recorded between included studies (*I²* = 98.8%, *p* < 0.001) [13,14,20,23–25,29,43,46,48,49,51,53,54,56,58–62] (Fig. 4).

**Fig. 4.**
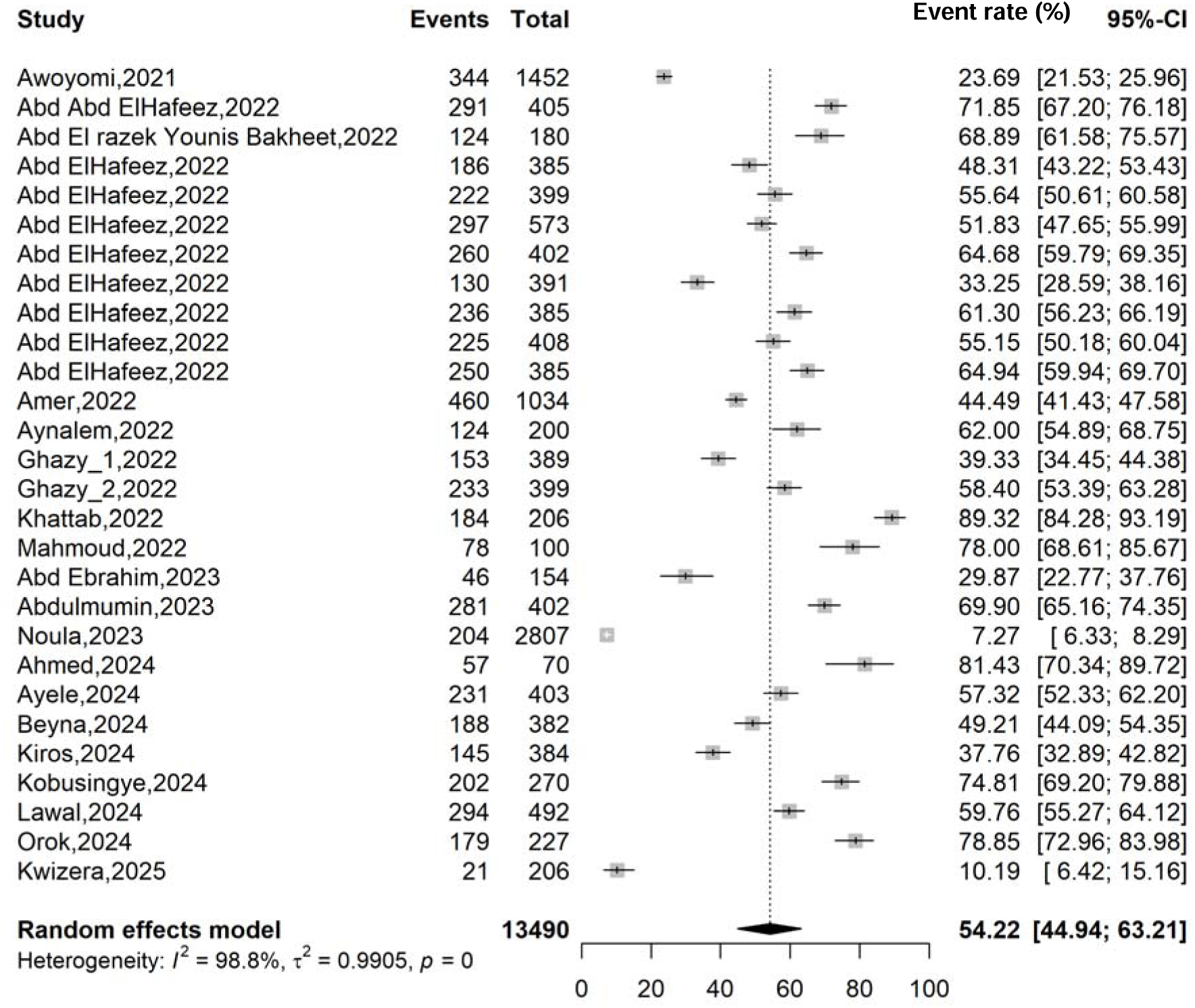
Forest plot displaying the pooled prevalence of positive perception/attitude toward mpox in Africa

The positive perception fluctuated over time, but remained stable at average level from 2021-2022 at 57.80% (95% CI: 49.18–65.97; n = 7,693; 17 reports) [13,25,29,43,46,48,49,51,53] and 2024-2025 at 55.80% (95% CI: 37.02–73.05; n = 2,434; 8 reports) [14,23,24,58–62], despite the non-significant (*p* = 0.362) drop in 2023 to 29.81% (95% CI: 8.09–67.20; 3 reports, n = 3,363)[20,54,56] . Significant disparities were observed by participant type (*p* < 0.001). Actually, pregnant women reported the highest positive perception at 81.43% (95% CI: 70.34–89.72; n = 70; 1 study) [58], followed by teachers at 68.89% (95% CI: 61.58–75.57; n = 180; 1 study) [46] and healthcare workers at 60.32% (95% CI: 53.50–66.77; n = 8,218; 21 reports) [13,14,24,25,29,48,49,51,53,54,59–61]. Conversely community health workers demonstrated the lowest positive perception at 10.19% (95% CI: 6.42–15.16; n = 206; 1 study) [62]. Positive perception was significantly (*p* < 0.001) most prevalent in Northern Africa, with a pooled proportion of 63.02% (95% CI: 48.84–75.25; n = 3,101; 8 reports) [25,29,46,49,51,53,56,58], closely followed by Southern Africa at 61.30% (95% CI: 56.23–66.19; n = 385; 1 study) [29]. Western and Eastern Africa showed moderate levels, with pooled proportions of 55.50% (95% CI: 41.23–68.91; n = 4,160; 8 reports) [24,29,43,48,54,59] and 50.84% (95% CI: 37.11–64.45; n = 3,037; 9 reports) [13,14,23,29,60–62] respectively. Conversely, Central Africa demonstrated a low pooled proportion of just 7.27% (95% CI: 6.33–8.29; n = 2,807; 1 study) [20] (Table 3 and Additional File 2, Supplementary Fig 1-8).

**Table 3.** Meta-regression analysis of pooled estimates of good mpox knowledge and positive perception prevalences in Africa.

| Estimate | Moderator | Univariate |  | Multivariate |  |
| --- | --- | --- | --- | --- | --- |
|  |  | Unadjusted coefficient (β) | p-value | Adjusted coefficient (β) | p-value |
| Knowledge |  |  |  |  |  |
| Study year | Year | 0.1835 | 0.3625 | 0.2770 | 0.203 |
| Design | Quasi-experimental vs. Cross-sectional | 2.4633 | 0.006 | 3.1962 | 0.001 |
| Participant | Healthcare worker vs. Others | 0.7807 | 0.033 | 0.6464 | 0.212 |
| Setting | Hospital vs. Others | 1.0695 | 0.002 | 0.6314 | 0.194 |
| WHO zone<sup>1</sup> | Western | Ref |  | Ref |  |
|  | Northern | -0.2894 | 0.553 | -0.6440 | 0.159 |
|  | Eastern | -0.1455 | 0.757 | -0.1980 | 0.669 |
|  | Southern | 1.3602 | 0.308 | 0.9142 | 0.433 |
|  | Central | -1.3356 | 0.168 | -1.0061 | 0.365 |
|  | Multi-region | -0.8223 | 0.537 | -0.8345 | 0.529 |
| WHO Afro<sup>2</sup> | Yes | Ref |  | - | - |
|  | No | -0.1907 | 0.661 | - | - |
|  | Multi-region | -0.7234 | 0.582 | - | - |
| Sampling method | Probabilistic vs. Non-probabilistic | 0.1595 | 0.699 | -0.2004 | 0.689 |
| Sample size | Number of participants | -0.0004 | 0.250 | 0.0002 | 0.652 |
| Perception |  |  |  |  |  |
| Study year | Year | -0.1071 | 0.583 | -0.0134 | 0.947 |
| Design | Quasi-experimental vs. Cross-sectional | 1.0289 | 0.169 | 1.7911 | 0.043 |
| Participant | Healthcare worker vs. Others | 1.0413 | 0.010 | 1.3386 | 0.024 |
| Setting | Hospital vs. Others | 0.5688 | 0.140 | -0.2705 | 0.546 |
| WHO zone<sup>1</sup> | Western | Ref |  | Ref |  |
|  | Northern | 0.3073 | 0.486 | 0.0501 | 0.907 |
|  | Eastern | -0.1862 | 0.672 | -0.0133 | 0.979 |
|  | Southern | 0.2396 | 0.802 | 0.0488 | 0.955 |
|  | Central | -2.7666 | 0.004 | 0.4722 | 0.791 |
| WHO Afro<sup>2</sup> | Yes vs. No | -0.5336 | 0.191 | - | - |
| Sampling method | Probabilistic vs. Non-probabilistic | -0.6435 | 0.1215 | -0.2745 | 0.570 |
| Sample size | Number of participants | -0.0012 | <0.001 | -0.0009 | 0.224 |

<sup>1</sup> Western: Algeria, Nigeria, and Senegal; Northern: Egypt, Libya, and Morocco; Eastern: Ethiopia, Kenya, Rwanda, Sudan, Tanzania, and Uganda; Southern: South Africa; Central: Cameroon; WHO: World Health Organization; <sup>2</sup> Redundant predictor dropped from the model;

The map of mpox positive perception in Africa showed significant disparities across countries (*p* < 0.001). It ranged from very high perception in countries like Egypt (64.43%, 95% CI: 48.76–77.52; n=2,528; 8 studies) and Algeria (71.85%, 95% CI: 67.20–76.18; n=405; 1 study) to critically low levels in Cameroon (7.27%, 95% CI: 6.33–8.29; n=2,807; 1 study) (Fig. 5).

**Fig. 5.**
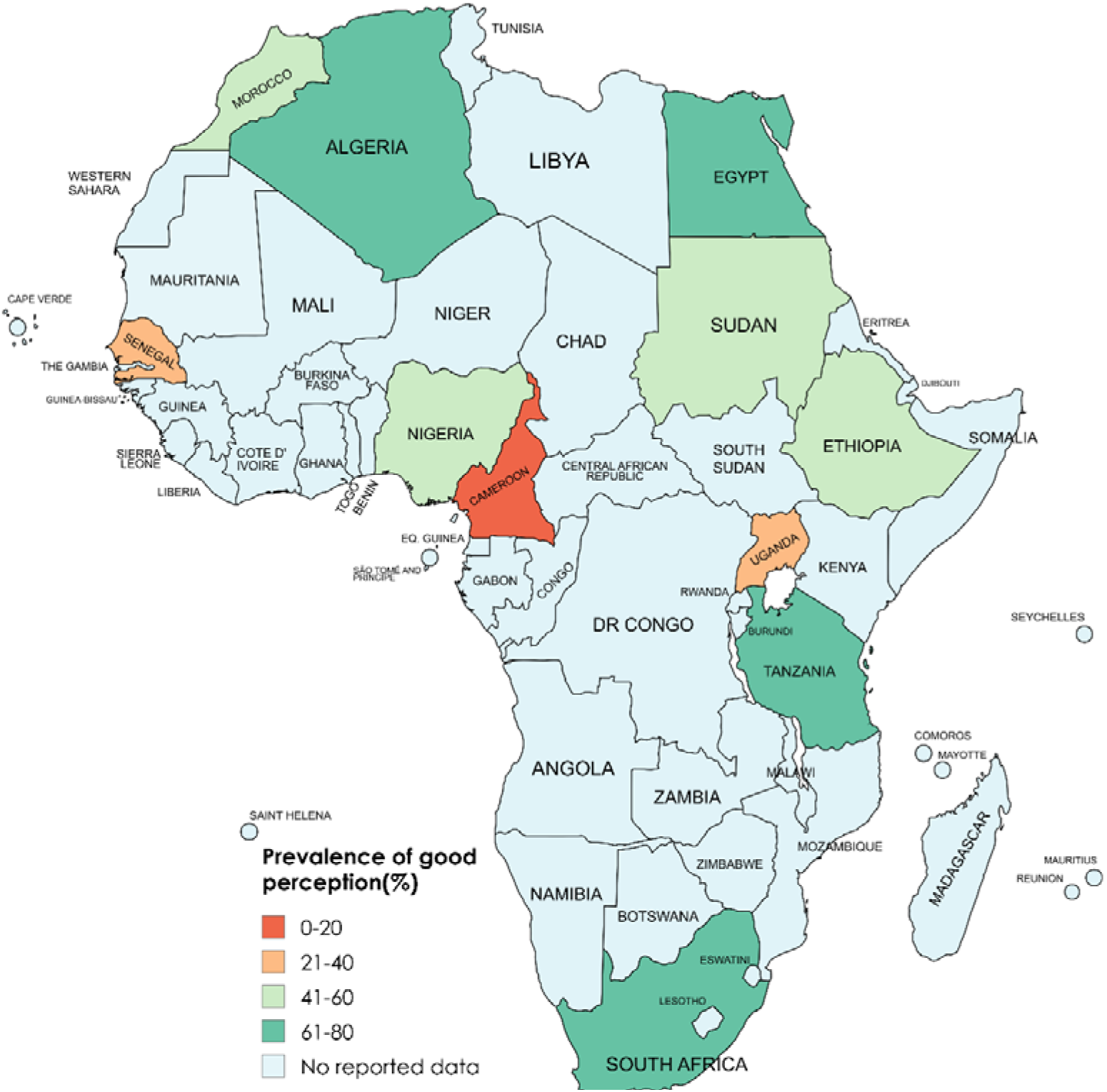
Map of the prevalence of positive mpox perception across African countries (Additional File 3, Supplementary Fig. 2)

### 3.5. Meta-regression analysis

The multivariate analysis revealed that the design of included studies was a significant sourced of heterogeneity of mpox knowledge estimates in Africa (β = 3.1962; *p* < 0.001). Moreover, studies measuring positive mpox perception were significantly influenced by study design (β = 1.7911; p < 0.043) and type of participants (β = 1.3386; *p* < 0.024) (Table 4).

**Table 4.**
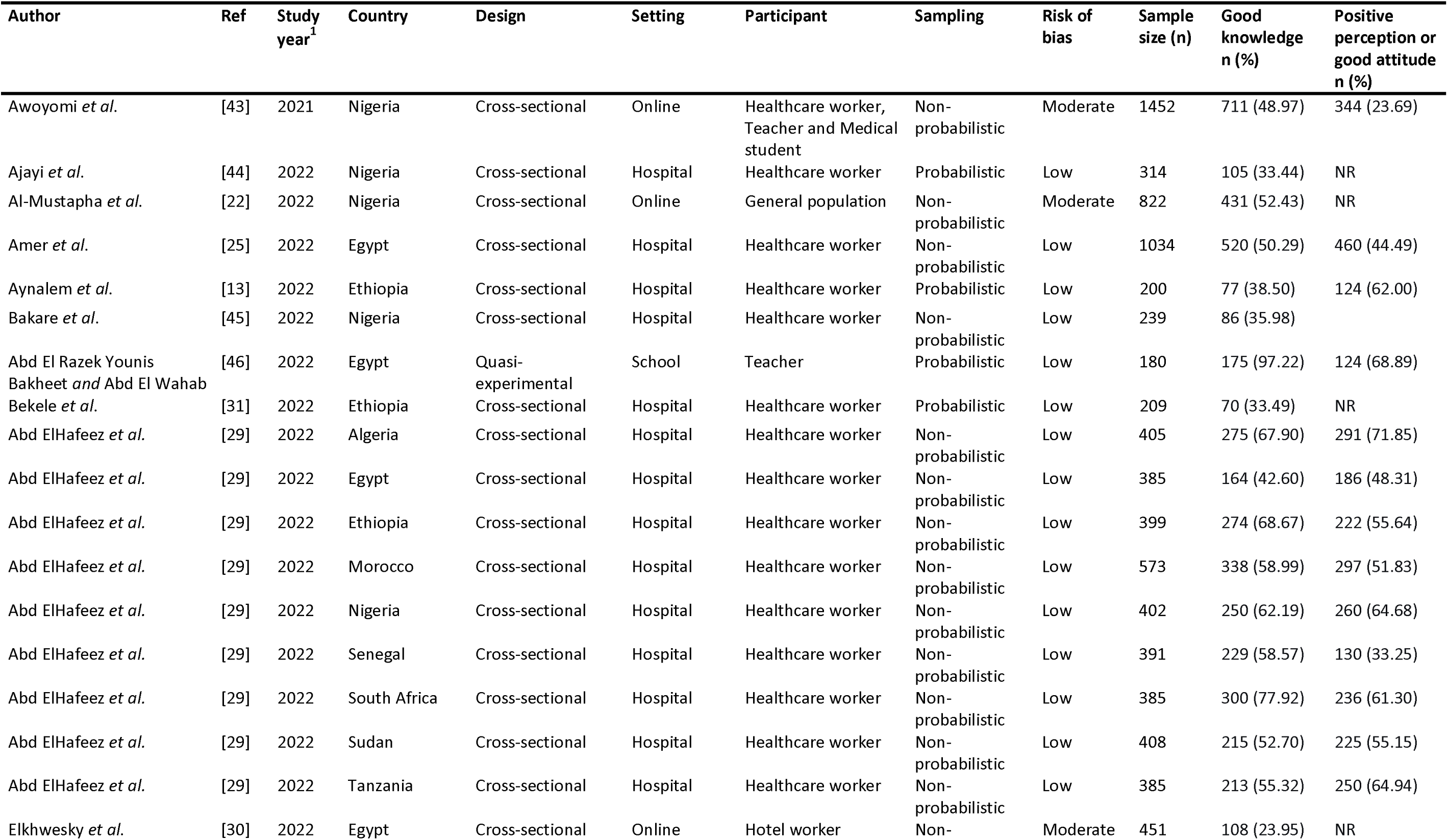
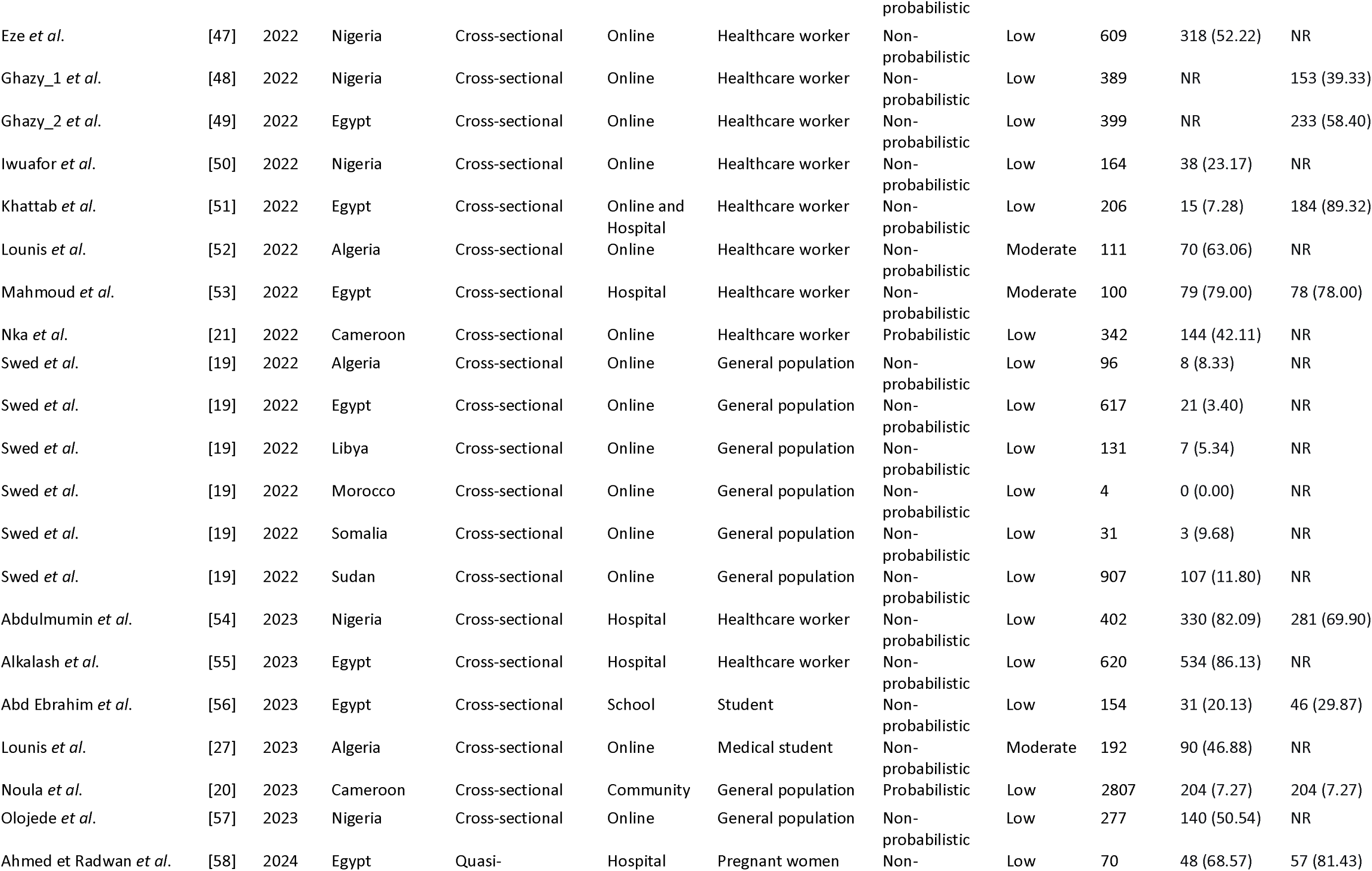
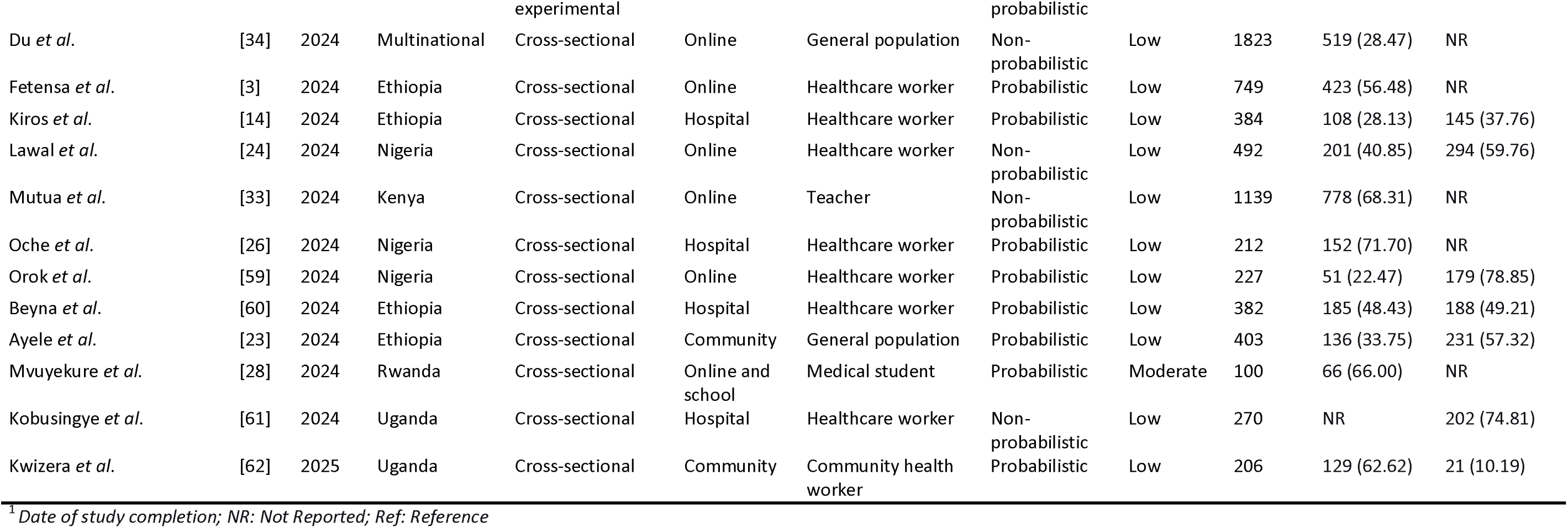
Characteristics of included studies.

### 3.6. Publication bias and sensitivity analysis

The funnel plot suggests no sign of publication bias for mpox knowledge pooled prevalence. In addition, the Egger’s (p = 0.896) and the Begg’s tests (p = 0.286) further confirmed this observation. No study significantly influences the pooled prevalence of mpox knowledge suggesting robustness of the pooled estimate (Additional File 1, supplementary Fig. 9 and 10).

Pooled prevalence estimates of positive mpox perception showed signs of publication bias (funnel plot asymmetry and test of Egger’s p = 0.010). The adjusted pooled estimate after performing the trim and fill analysis (eight hypothetical studies added) was 42.91% whose 95%CI 33.38-53.01 overlapped with the primary pooled estimate of 54.22% (95%CI: 44.94-63.21) suggesting no significant difference between the two pooled estimates. The sensitivity analysis revealed that the recalculated proportion remained within a narrow range of 40% to 52% regardless of which individual study was omitted, confirming that the overall meta-analysis result was not disproportionately influenced by any single study (Additional File 2, Supplementary Fig 9-11).

### 3.7. Systematic synthesis of determinants

#### 3.7.1. Socioprofessional characteristics

##### Gender

Male participants were found to have significantly higher knowledge scores about mpox than females in some studies [22,23]. Conversely, male participants reported lower mpox vaccine hesitancy and more positive attitudes toward vaccination compared to females [3,33,48].

##### Age

Younger adults (e.g., <35 years) were associated with better knowledge of mpox [22,23]. In contrast, older healthcare workers demonstrated a more positive attitude toward mpox vaccination compared to their younger colleagues [3,48].

##### Education

Higher educational attainment (university level) was a socio-demographic predictor of superior knowledge and more positive health attitudes regarding mpox disease [19,23,24,29,51,55].

##### Profession

As compared to the general population, healthcare workers and teachers consistently demonstrate significantly higher levels of knowledge and more positive attitude [3,13,25,31,44,45,47–49,52,54,55,59,60].

##### Specific healthcare profession

Physicians ranked highest, consistently exceeding the knowledge levels of nurses, midwives, and allied health staff [13,31,44,47,59]. This knowledge advantage corresponded with more positive attitudes and a greater intention to be vaccinated compared to other professional groups [3,48].

##### Localization

Residing in an urban setting was linked to better knowledge [22,23].

##### Workplace

Healthcare workers employed in tertiary or specialized hospitals reported higher knowledge than those in primary healthcare centers, possibly due to better access to resources, training, and a more complex case load [21,45].

##### Clinical experience and work in high-risk departments

Healthcare workers with more years of experience, particularly those who had managed infectious diseases or worked in departments like infectious diseases, emergency, or dermatology, had superior knowledge [13,28,44,47,50].

##### Being a medical or life sciences student

Students pursuing medical, nursing, or life sciences programs demonstrated significantly higher levels of knowledge compared to their peers in other academic discipline [27,29].

#### 3.7.2. Non-socioprofessional characteristics

##### Source of information

Sourcing information from credible channels such as medical journals, official health guidelines, and academic training was linked to more accurate knowledge [19,22,24,29,44,55]. This subsequently reflected to greater trust in official recommendations and a more positive attitude toward public health measures [34,48,49].

##### Good mpox knowledge

Individuals with a better understanding of mpox transmission, symptoms, and prevention had more positive attitudes [3,13,25,27,49,52,60].

##### Formal training and educational interventions

Attending targeted mpox educational sessions, workshops, or structured training programs is a powerful, modifiable factor that directly leads to significant improvements in both knowledge scores and positive attitudes [29,51,53,58].

##### Perceived susceptibility and disease severity

The perception of personal susceptibility to mpox infection and the perceived severity of the disease were both correlated with a more positive attitude toward adopting protective behaviors, such as vaccination and follow hygiene measures [3,48].

##### Media exposure

Frequent exposure to news and information about mpox through television, radio, or the internet was positively correlated with good knowledge levels [22,28].

## 4. Discussion

This systematic review and meta-analysis across Africa from 2021 to 2025 provides a detailed understanding of Mpox knowledge and attitudes. Despite Africa’s long-standing experience with mpox, the overall knowledge level remains low, and attitudes are only average. These findings highlight ongoing gaps in emergency public health preparedness and the need for more robust and context-specific education and communication strategies.

### 4.1. Knowledge of mpox

In our study, the pooled prevalence of good mpox knowledge was below average (43%), despite the disease’s longstanding presence in Africa, indicating significant gaps in awareness among African populations. This gap in knowledge likely stems from the historical neglect of mpox and other zoonotic diseases within health education, limited public health messaging, and the overwhelming focus on other high-burden diseases like HIV, tuberculosis, and malaria in African health systems [63,64]. Due to their frontline role in detection and response, healthcare workers generally demonstrated better knowledge than the general public. However, our finding demonstrated that their overall understanding remained insufficient as reported elsewhere [13,14]. This is likely due to insufficient mpox-specific training, limited access to updated guidelines, and weak continuing professional development systems, especially in primary or rural healthcare settings across Africa [63,65]. These results show the need for fair resource distribution, ongoing training for healthcare workers, and more health education for everyone.

Teachers and medical students showed the highest knowledge of mpox in our analysis. This is likely because of their education, access to academic resources, structured learning, and digital information [27,46]. On the other hand, the general public had very low awareness in countries like Cameroon, Ethiopia, Nigeria, and Somalia. This may be because they rely on informal sources and have limited access to reliable health information [20,22,23].

Additionally, large regional disparities were evident. Southern Africa had the highest knowledge levels, consistent with stronger health infrastructures and greater access to training opportunities [29]. On the other hand, regions like Central Africa, although historically the center of mpox outbreaks, showed some of the lowest awareness levels in our analysis. This difference could be due to long-standing weaknesses in health communication, diagnostic limitations, and ongoing underinvestment in surveillance and outbreak preparedness [10,12]. Lastly, our analysis revealed variation across countries; for instance, Libya and Somalia had very low knowledge levels, while Kenya, South Africa, Rwanda, and Uganda showed relatively higher knowledge among surveyed populations [28,33,62]. The variation might be due to differences in healthcare systems, information sources, and educational levels.

### 4.2. Attitudes toward mpox

Our analysis shows that positive perceptions of mpox were average (54%) but varied across different groups. People with greater knowledge generally have a more favorable view of mpox prevention and vaccination, highlighting the strong role of health literacy in shaping risk-prevention actions and perceptions [3,10,21]. Other groups in our study who displayed positive attitudes included teachers and pregnant women. Their favorable attitudes are likely due to greater exposure to health information or participation in structured educational programs [46]. In contrast, community health workers and those relying on informal information sources showed less confidence and greater hesitancy, likely due to misinformation, unclear risk messaging, and a low sense of personal risk [45]. These findings are consistent with previous reports from the Middle East, North Africa, and globally, which document misinformation and low perceived risk as barriers to positive attitudes [10,19,34,48], highlighting the broader challenge of maintaining accurate public understanding during emerging infectious disease outbreaks.

### 4.3. Variation in knowledge and attitudes

Several factors were linked to differences in knowledge and attitudes in our analysis. These differences reflect not only sociodemographic variations but also fundamental structural and informational inequalities. Populations with reliable access to accurate information, such as healthcare workers in tertiary facilities, individuals with clinical exposure, and those who received formal training, showed stronger knowledge and more positive attitudes [23,27,33]. The ongoing success of educational interventions highlights the value of structured learning in increasing awareness and influencing attitudes.

On the other hand, people in rural areas, those who rely on social media, and groups with little health education had lower knowledge and weaker attitudes. This is likely because they have less access to health information, face more misinformation, and feel less at risk [20,21]. These trends fit with known behavioral models, which show that knowledge, feeling at risk, and trust in health authorities all influence attitudes and actions [3].

### 4.4. Alignment with global evidence

The data from Africa aligns with global trends. Jahromi *et al.* found in a worldwide review that about half of healthcare workers do not know enough about mpox, especially about diagnosis, prevention, and vaccination [38]. Likewise, León-Figueroa *et al.* also found poor to moderate understanding and very mixed attitudes, often affected by misinformation and low risk perception [18]. Global patterns of hesitancy about mpox vaccination and other preventive strategies are similar to those in Africa, suggesting that preparedness challenges are not limited to one region. These results confirm that the issues affecting mpox knowledge and attitudes are part of a wider global problem in communicating about new infectious diseases, not just an African issue.

## 5. Limitations

There are several limitations to this review that should be kept in mind when looking at the findings. Many studies used self-administered online questionnaires, which could lead to selection bias and may overrepresent people with internet access or higher education. The studies also varied widely in their settings, populations, measurement tools, and how they defined “good knowledge” or “positive attitude.” In addition, some countries and subregions in Africa did not have published data, which limits how well the results represent the whole continent. Many studies used non-probabilistic sampling, which may affect how generalizable the findings are. Study reports published in languages other than French and English might not have been identified. Finally, publication bias could have influenced the results for attitudes, but sensitivity analyses showed this had little effect on the overall estimates.

## 6. Conclusions

Overall, knowledge and attitudes about mpox in Africa are suboptimal and differ by region. Healthcare workers know more than the general public, but there are still big gaps, reflecting issues with training, communication, and preparedness. These findings highlight the urgent need for better education, targeted messages, and more support for areas where mpox is common, to improve readiness for mpox and other new diseases both in Africa and worldwide.

## Supporting information

Addition files 2

## Data Availability

All data produced in the present work are contained in the manuscript

## 7. Abbreviations

*CI*: Confidence Interval
*MeSH*: Medical Subject Headings
*PRISMA*: Preferred Reporting Items for Systematic Reviews and Meta-Analysis

## 8. Declarations

### Ethical approval statement and consent to participate

Not applicable

### Consent for publication

Not applicable.

### Availability of data and materials

The sources of data supporting this systematic review are available in the reference. All data generated or analyzed during this study are included in this published article and supplemental material.

### Author contributions

F.Z.L.C. conceived the original idea of the study. F.Z.L.C. conducted the literature search. F.Z.L.C., R.T, A.N., C.A., and C.T.A. selected the studies, extracted the relevant information, performed quality assessment and synthesized the data. F.Z.L.C. performed the analyses and wrote the first draft of the manuscript. All authors critically reviewed and revised successive drafts of the manuscript. All authors read and approved the final manuscript.

## Acknowledgment

None.

