## Supplementary material for "Mpox knowledge and perception in Africa: a systematic review and meta-analysis": Addition files 2

**Subgroup analysis**

**Study period**

**Prevalence of positive perception (%)**

**Event rate (%)**

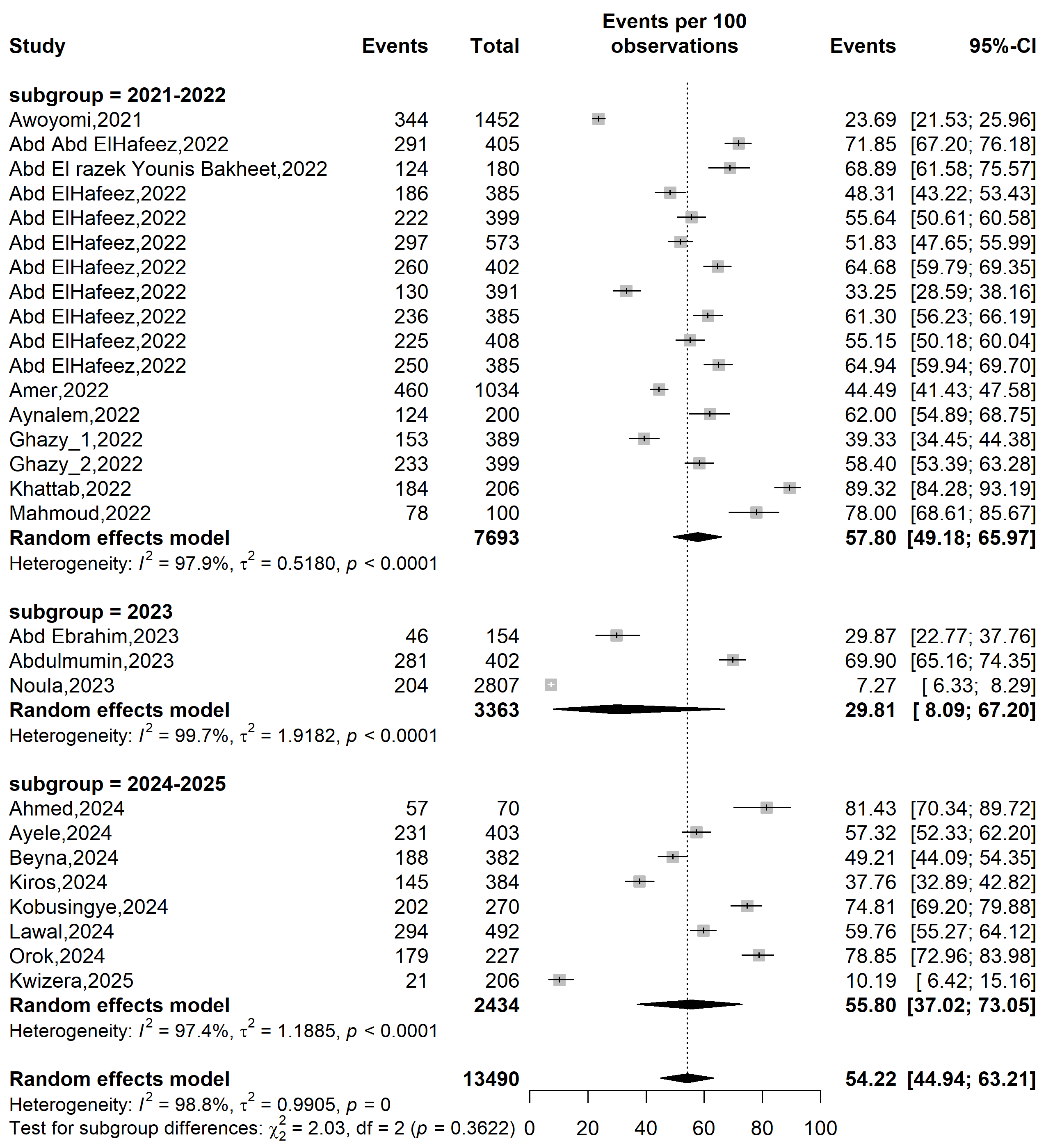

**Supplementary Fig. 1** Forest plot displaying the prevalence of positive perception of mpox in Africa according the study period

**Country**

**Prevalence of positive perception (%)**

**Event rate (%)**

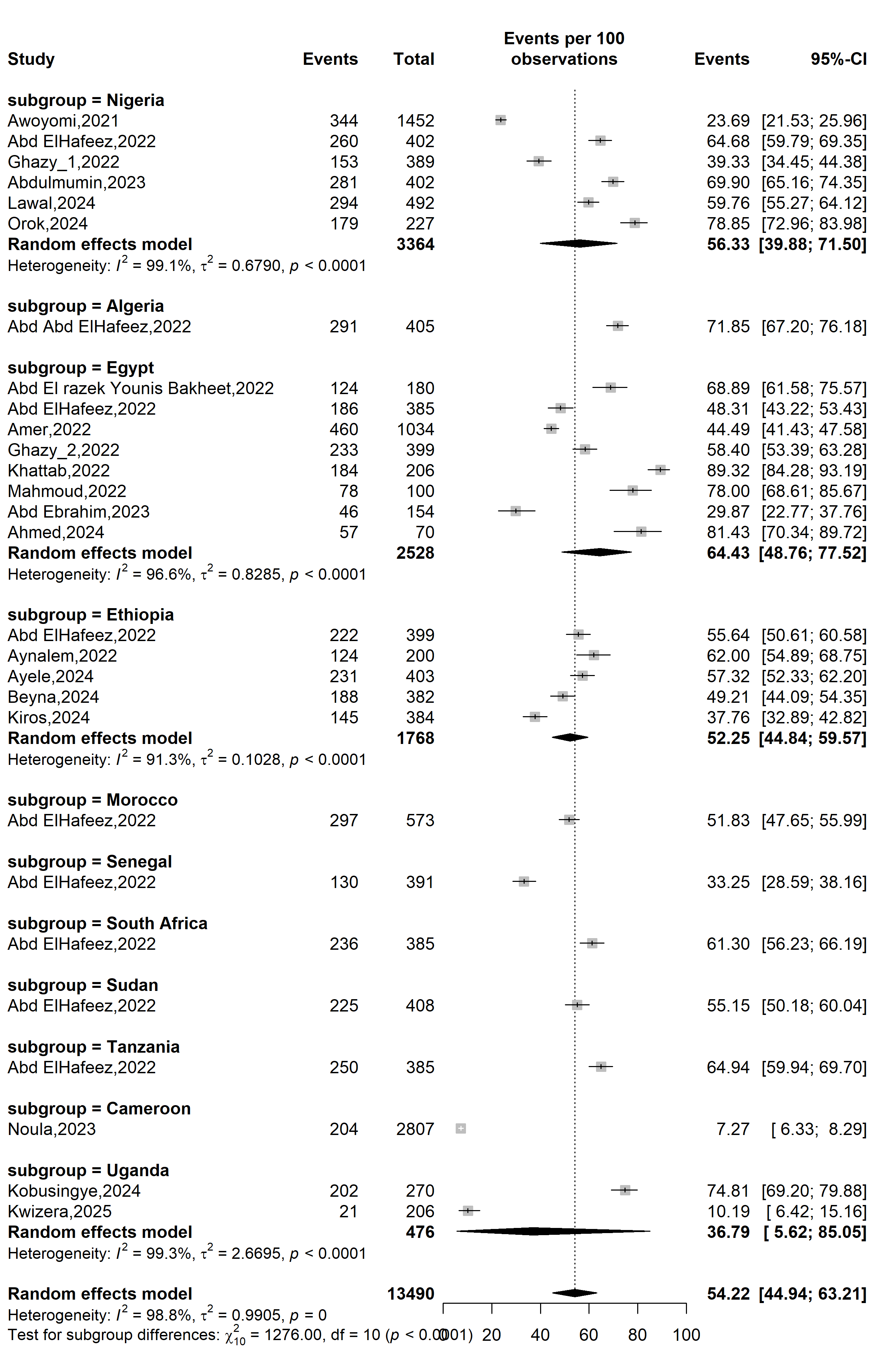

**Supplementary Fig. 2** Forest plot displaying the prevalence of positive perception of mpox in Africa by countries

**Participant**

**Prevalence of positive perception (%)**

**Event rate (%)**

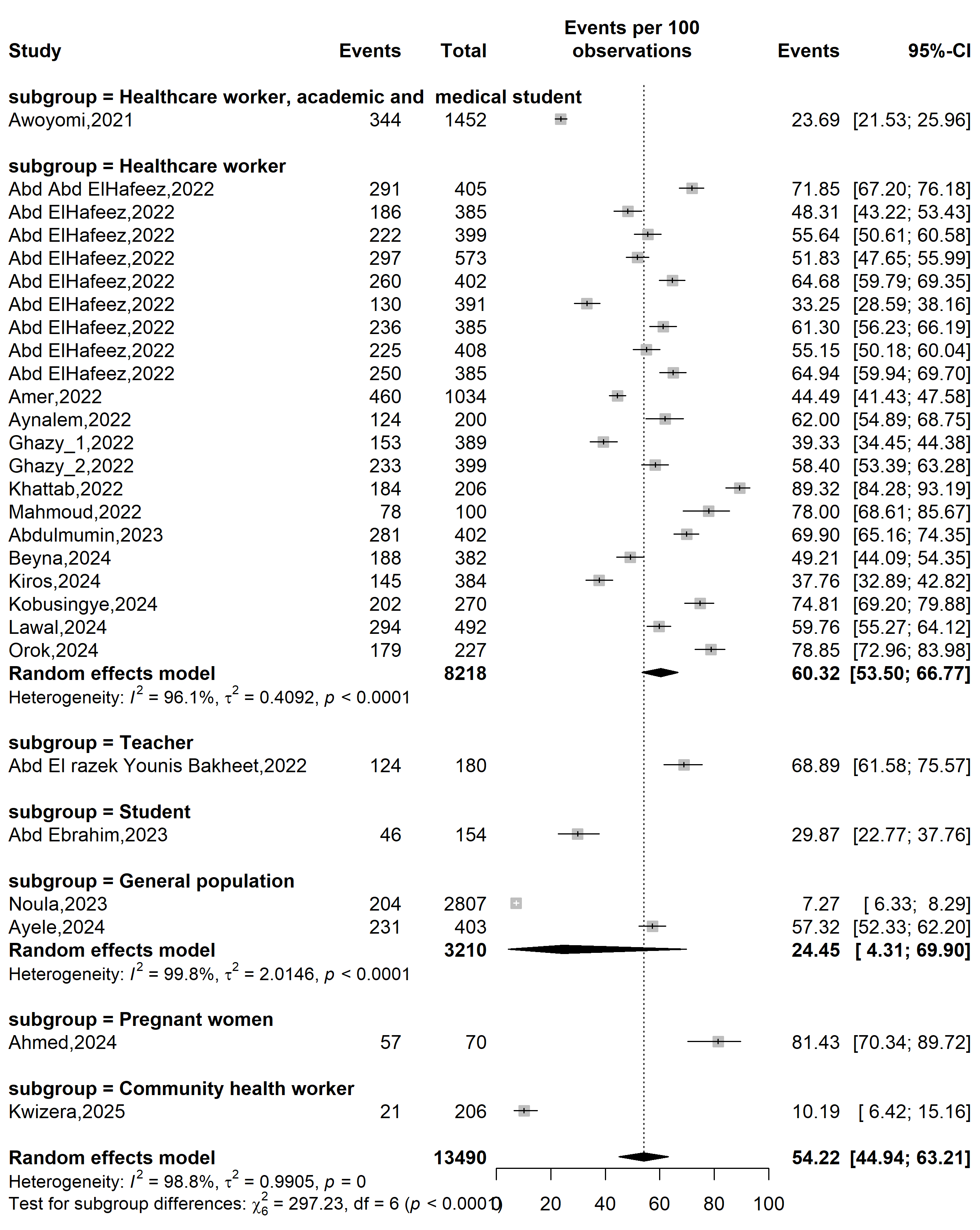

**Supplementary Fig. 3** Forest plot displaying the prevalence of positive perception of mpox in Africa by type of participants

**Study design**

**Prevalence of positive perception (%)**

**Event rate (%)**

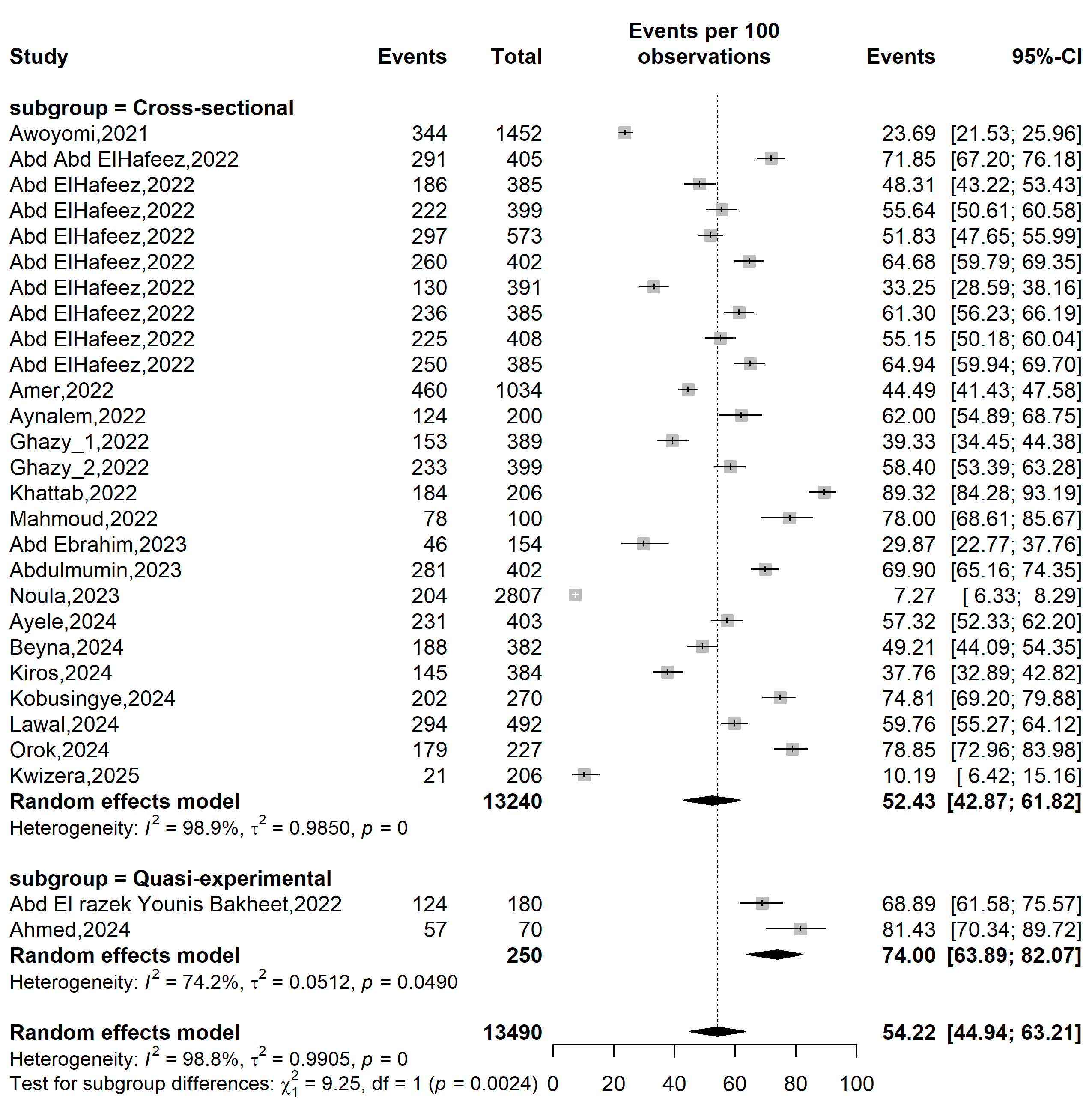

**Supplementary Fig. 4** Forest plot displaying the prevalence of positive perception of mpox in Africa according the study design

**Study setting**

**Prevalence of positive perception (%)**

**Event rate (%)**

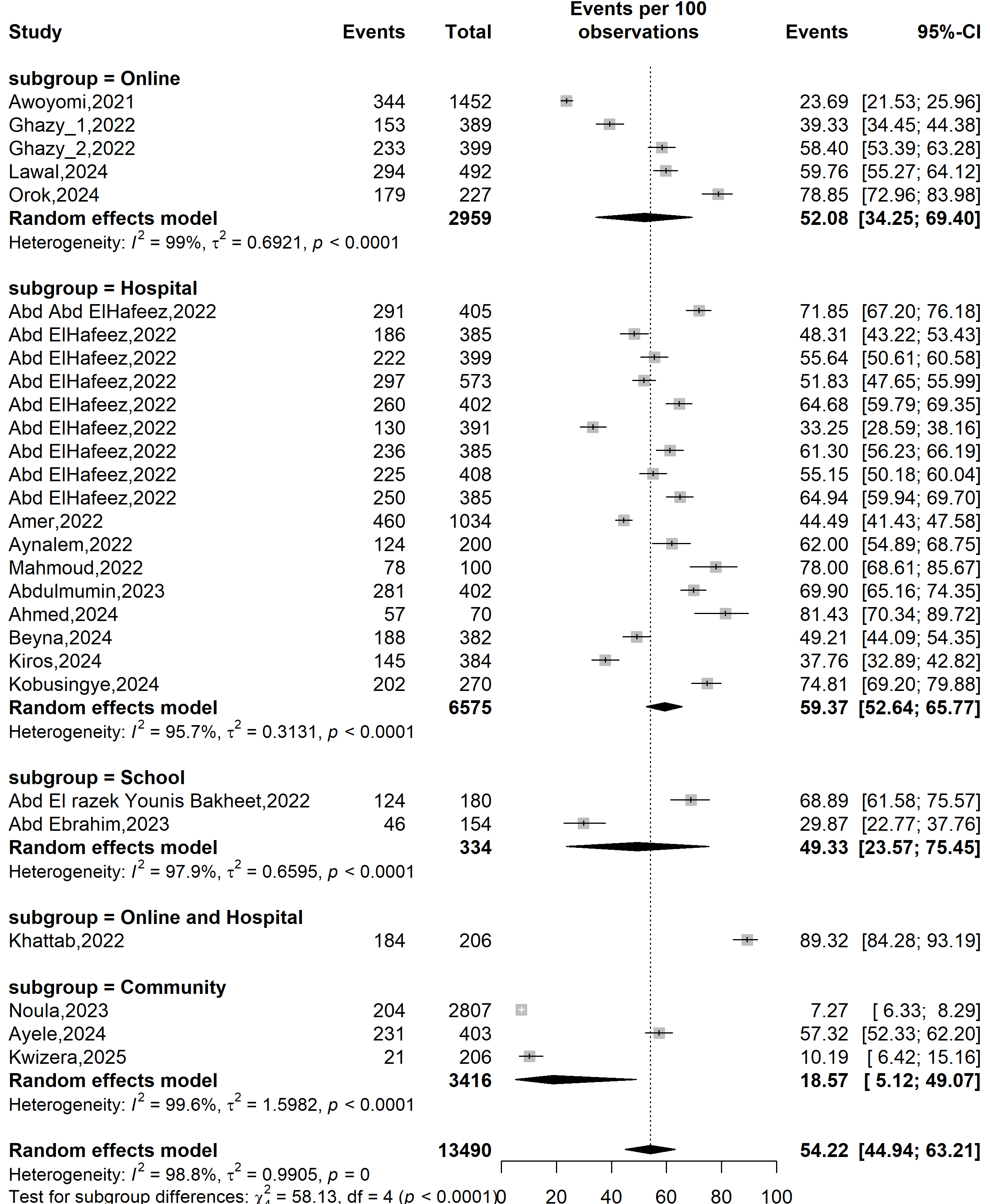

**Supplementary Fig. 5** Forest plot displaying the prevalence of positive perception of mpox in Africa according the study setting

**WHO region**

**Prevalence of positive perception (%)**

**Event rate (%)**

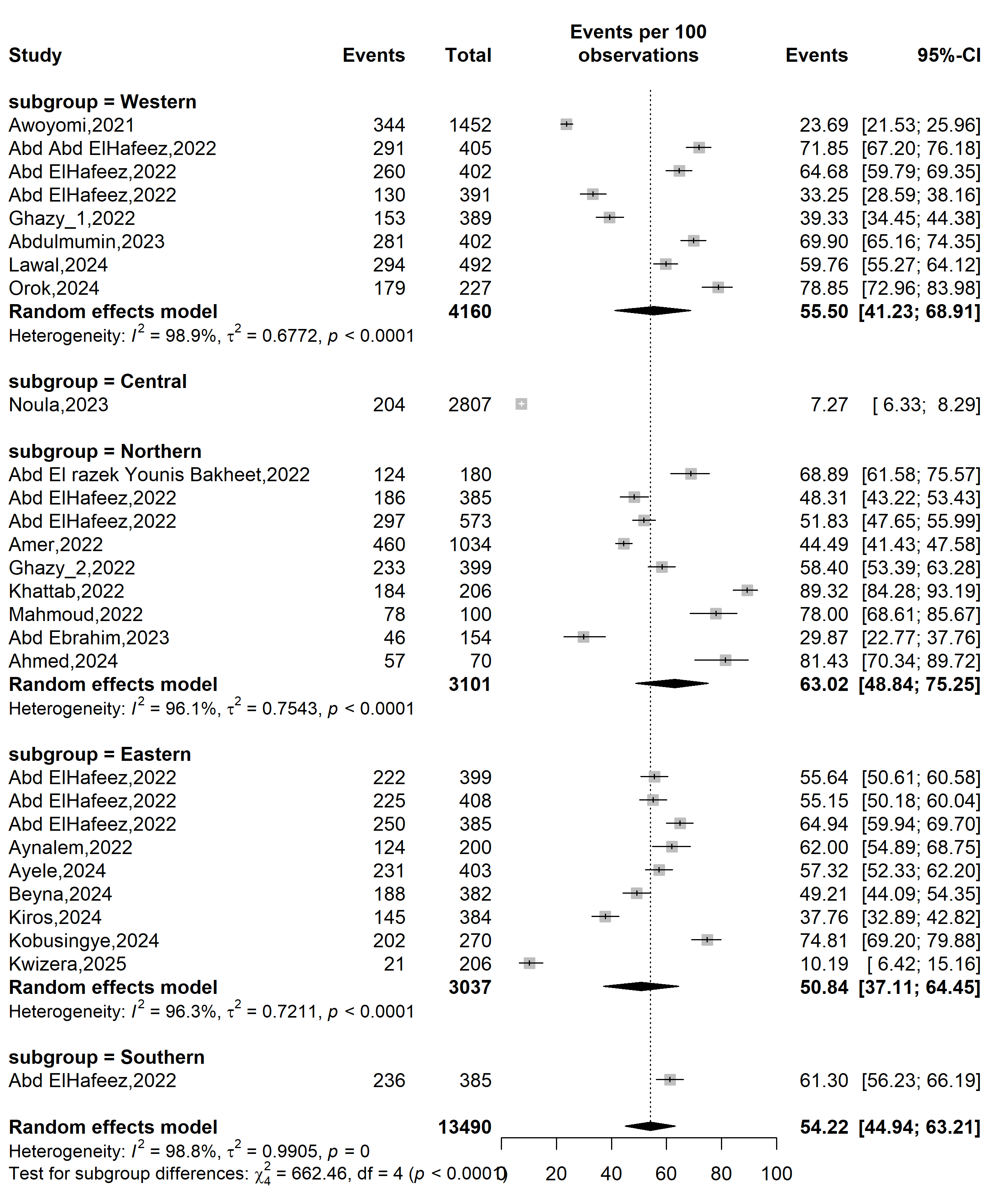

**Supplementary Fig. 6** Forest plot displaying the prevalence of positive perception of mpox in Africa by WHO Afro regions

**Event rate (%)**

**Prevalence of positive perception (%)**

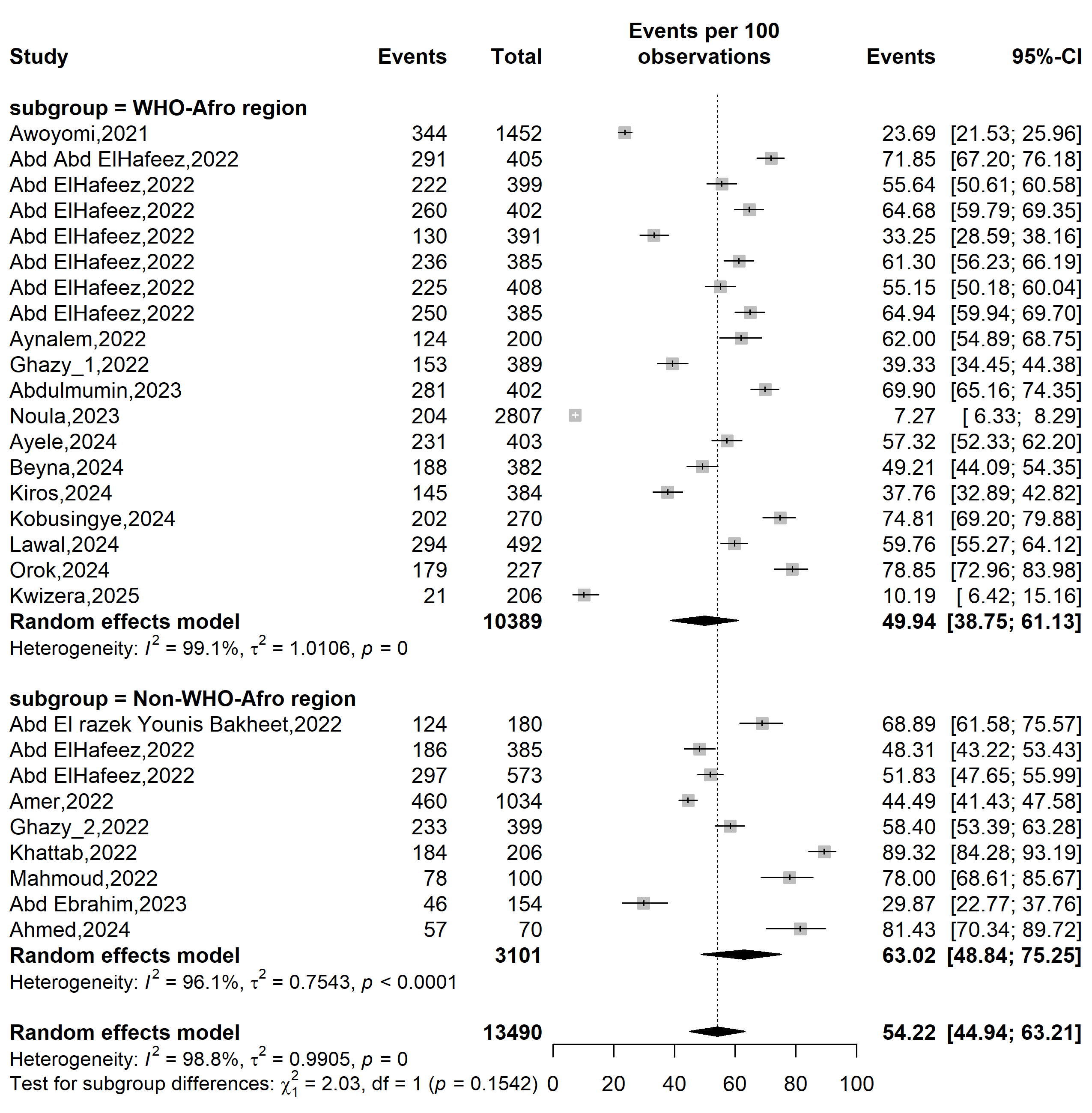

**Supplementary Fig. 7** Forest plot displaying the prevalence of positive perception of mpox in Africa by WHO regions

**Sampling**

**Prevalence of positive perception (%)**

**Event rate (%)**

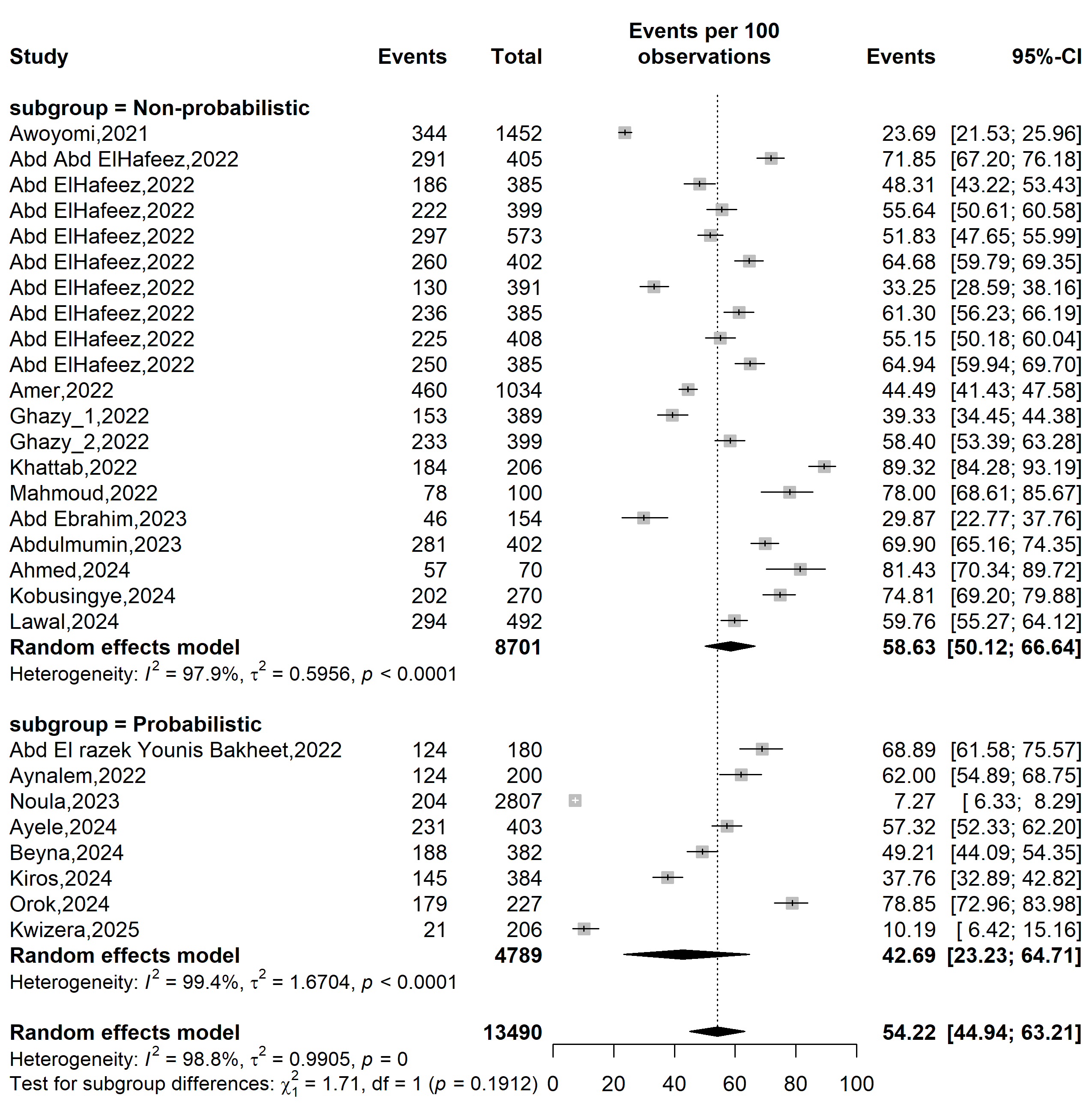

**Supplementary Fig. 8** Forest plot displaying the prevalence of positive perception of mpox in Africa by sampling method

**Publication bias assessment**

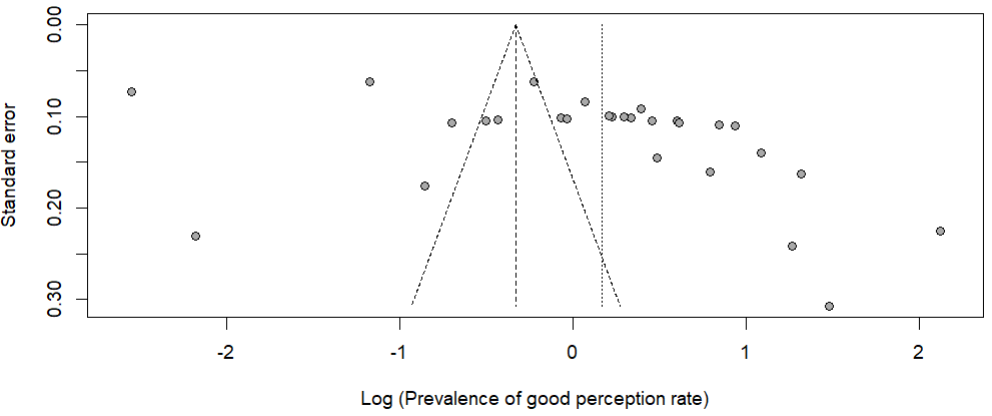

Egger’s test *p*-value = 0.010

Begg’s test *p*-value = 0.057

**Supplementary Fig. 9** Funnel plot displaying the pseudo 95% confidence limits and tests assessing the publication bias of studies included

**Trim and fill analysis**

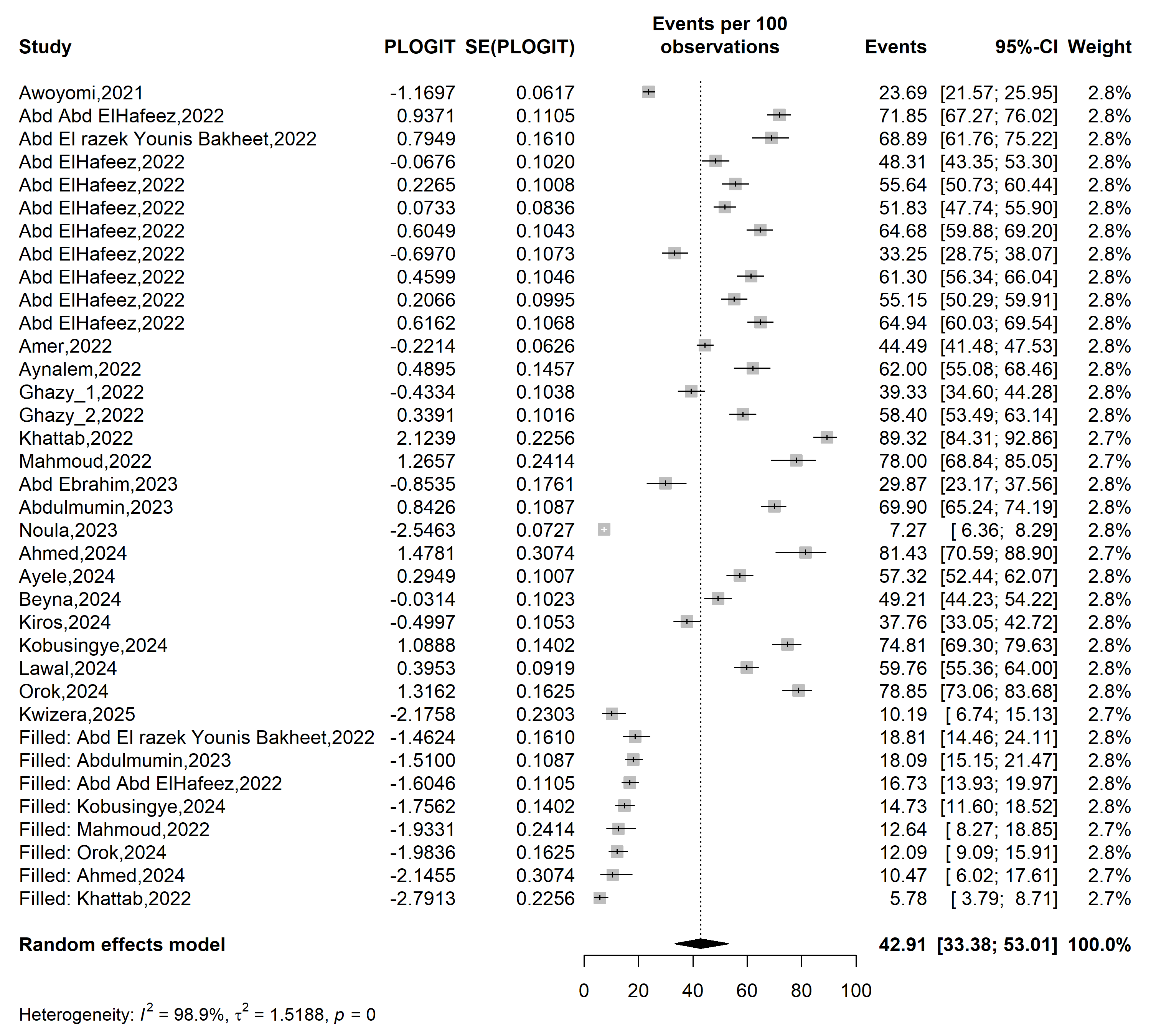

**Prevalence of positive perception (%)**

**Event rate (%)**

**Supplementary Fig. 10** Trim and fill plot of the prevalence of positive mpox perception in Africa

**Sensitivity analysis**

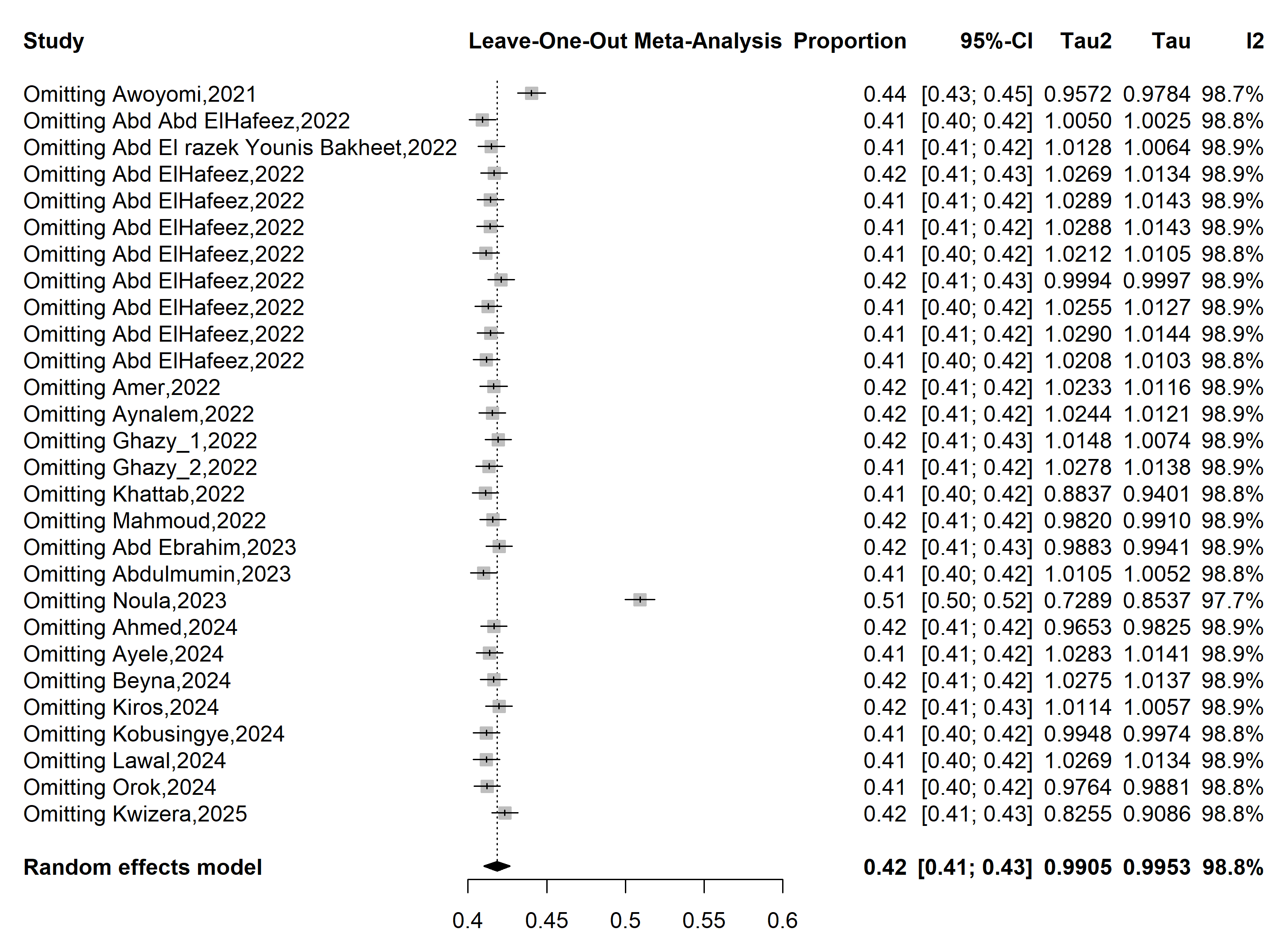

**Supplementary Fig. 11** Sensitivity analysis of the prevalence of positive perception of mpox in Africa
